# Joint analysis reveals shared autoimmune disease associations and identifies common mechanisms

**DOI:** 10.1101/2021.05.13.21257044

**Authors:** Matthew R Lincoln, Noah Connally, Pierre-Paul Axisa, Christiane Gasperi, Mitja Mitrovic, David van Heel, Cisca Wijmenga, Sebo Withoff, Iris H Jonkers, Leonid Padyukov, International Multiple Sclerosis Genetics Consortium, Stephen S Rich, Robert R Graham, Patrick M Gaffney, Carl D Langefeld, David A Hafler, Sung Chun, Shamil R Sunayev, Chris Cotsapas

**Affiliations:** Department of Neurology, Yale School of Medicine, New Haven, CT; Blizard Institute, Queen Mary University of London, London, UK; Department of Genetics, University Medical Center Groningen, University of Groningen, Groningen, The Netherlands; Division of Rheumatology, Department of Medicine, Karolinska Institutet and Karolinska University Hospital, Stockholm, Sweden; Center for Public Health Genomics, University of Virginia, Charlottesville, VA; Department of Public Health Sciences, University of Virginia, Charlottesville, VA; Maze Therapeutics, South San Francisco, CA; Genentech, South San Francisco, CA; Genes and Human Disease Research Program, Oklahoma Medical Research Foundation, Oklahoma City, OK; Department of Biostatistics and Data Science, Wake Forest School of Medicine, Winston-Salem, NC; Center for Precision Medicine, Wake Forest School of Medicine, Winston-Salem, NC; Division of Genetics, Brigham and Women’s Hospital, Boston, MA; Department of Biomedical Informatics, Harvard Medical School, Boston, MA; Department of Genetics, Yale School of Medicine, New Haven, CT

## Abstract

Autoimmune and inflammatory diseases are polygenic disorders of the immune system. Many genomic loci harbor risk alleles for several diseases, but the limited resolution of genetic mapping prevents determining if the same allele is responsible, indicating a shared underlying mechanism. Using a collection of 129,058 cases and controls across six diseases, we show that ∼40% of overlapping associations are due to the same allele. We improve fine-mapping resolution for shared alleles two-fold by combining cases and controls across diseases, allowing us to identify more eQTLs driven by the shared alleles. The patterns of sharing indicate widespread shared mechanisms, but not a single global autoimmune mechanism. Our approach can be applied to any set of traits, and is particularly valuable as sample collections become depleted.

**One sentence summary:** Genetic mapping in autoimmune diseases increases genetic mapping resolution and reveals shared mechanisms

## Main Text

Autoimmune and inflammatory diseases are a heterogeneous group of disorders, where activation of both the adaptive and innate immune system coupled with loss of self-tolerance leads to target tissue destruction (*1*). These diseases are heritable, and genome-wide association studies (GWAS) have identified hundreds of susceptibility loci, confirming their polygenic nature (*2, 3*). Like other complex disease risk traits, heritability is strongly enriched in gene regulatory regions active in specific cell populations (*4–6*), suggesting risk is mediated to a large extent by altering gene expression in specific cell types under specific conditions. These diseases are also comorbid (*7, 8*), with dual diagnoses being more frequent in individuals than expected by chance, and multiple diseases aggregating in families (*9*). We and others have shown that many genetic loci harbor risk variants for multiple autoimmune diseases (*10–12*), suggesting that comorbidity may be due to shared genetic liability and, hence, shared mechanisms of disease.

Instances of pleiotropy, where the same variant influences risk to more than one disease, would by definition point to a shared molecular effect, and thus a shared mechanism. The limited resolution of genetic mapping has made it difficult to distinguish such cases from situations where distinct genetic variants in the same locus mediate risk to different diseases. This limited resolution restricts our ability to uncover shared pathogenic mechanisms, understand why some modulating immune functions can increase risk to one disease whilst decreasing risk to others, or make inferences about the origins of these diseases and their different prevalence rates around the world.

An important driver of the limited resolution of genetic mapping is disease cohort sample size (*13*). Currently available disease cohorts, most of which have been extensively studied already, are the result of decades-long international recruitment efforts. Meaningful increases in sample size are thus difficult to envision in the immediate future. An alternative way to increase sample size, and thus genetic mapping resolution, would be to jointly analyze cohorts across diseases. This would also reveal shared pathogenic mechanisms. In conventional meta-analyses of cohorts with the same disease, we assume that any associations are shared across strata; we cannot make this assumption across diseases. It is thus crucial to ensure that the same allele drives risk to two or more diseases, rather than separate alleles in the same genomic locus.

Here, we first show substantial genome-wide shared heritability between autoimmune and inflammatory diseases (Fig. S1). We then look at 224 instances where genetic associations to multiple diseases occur in the same genomic region, and show that 41.5% of these observed associations are due to pleiotropic variants, with the remainder being due to different alleles in the region. When we combine cases and controls across diseases to map each shared association, we increase fine-mapping resolution two-fold on average. Further, this increase in resolution reveals new disease risk variants that alter gene expression in immune cell subtypes. Comorbidity is widespread between diseases of all organ systems, and sample sizes are limited, so this strategy is widely applicable beyond the immune-mediated diseases. Thus, this approach to careful dissection of shared effects can reveal mechanisms that are common across diseases and pinpoint key genes driving shared biology.

We first assessed the evidence for genome-wide shared heritability between 19 autoimmune and inflammatory diseases from GWAS summary data. After quality control, we used LD score regression (*14*) to estimate heritability (*h*_*g*_^*2*^) for each trait (Fig. S2A). We found that 13/19 diseases had sufficient heritability captured by common variants to make these comparisons meaningful (*Z*-score > 4) (*15*), so we restricted our analysis to this subset. We then calculated the proportion of shared heritability between each pair of diseases, again using LD score regression, which is robust to sample overlaps between cohorts (*15*). We found a broad pattern of shared heritability (Fig. 1), with the strongest overlaps (0.63 ≤ *r*_*g*_ ≤ 0.92) between IBD and its subtypes, Crohn disease and ulcerative colitis; these are known to share the majority, but not the entirety, of their genetic architecture (*16*). We also observed strong correlations between atopic dermatitis, asthma and allergic traits (0.51 ≤ *r*_*g*_ ≤ 0.91), which may represent a shared basis for atopic inflammatory disease. We saw a strong correlation between systemic sclerosis and systemic lupus erythematosus, which were also correlated with primary biliary cirrhosis (0.42 ≤ *r*_*g*_ ≤ 0.86). In line with our previous findings (*10, 17*), these results indicate that autoimmune and inflammatory diseases share a substantial portion of genetic risk factors, even when accounting for the major histocompatibility locus (MHC), where overlapping haplotypes confer risk to different autoimmune and inflammatory diseases (*18*). Overall, this suggests that some mechanisms are common between sets of diseases, but we find no evidence of universal sharing indicative of a large core autoimmune susceptibility component (*10, 17*).

**Fig. 1:**
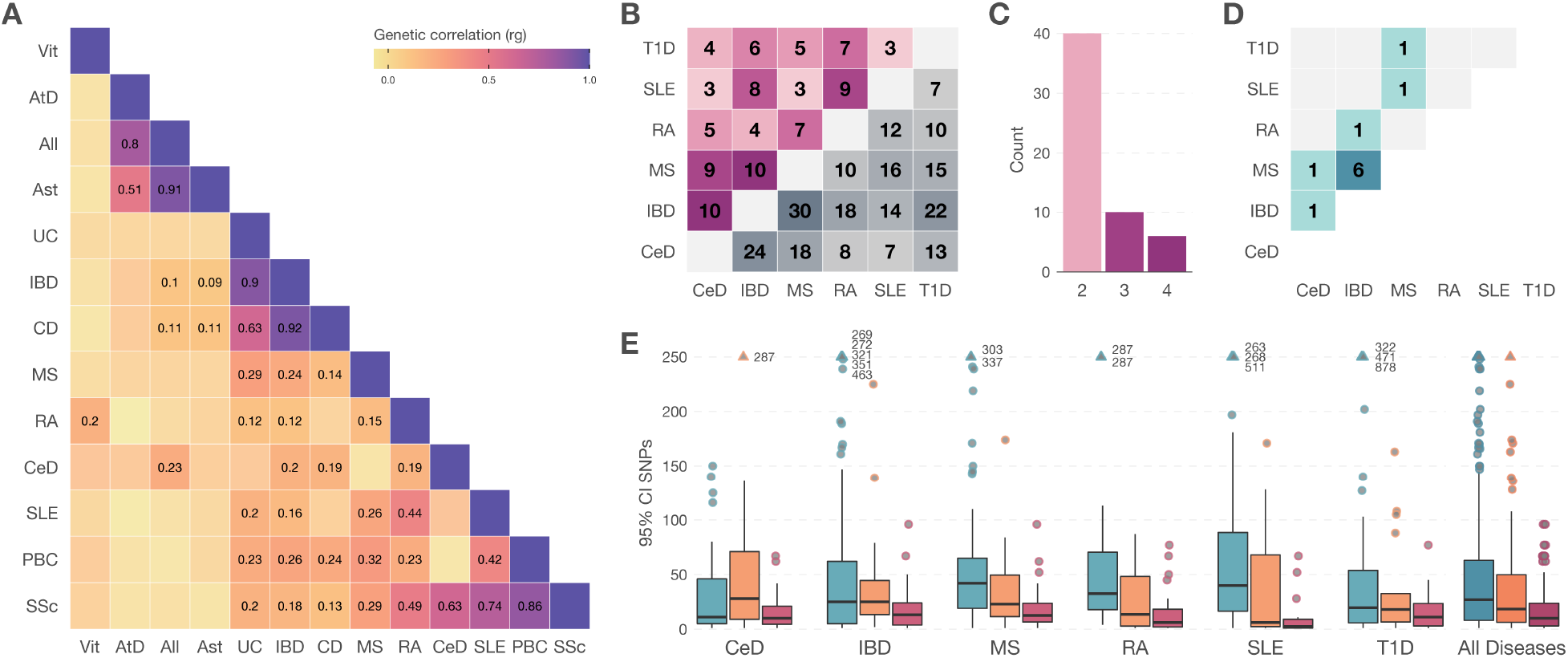
Joint analysis of shared autoimmune disease risk alleles improves fine-mapping two-fold. (A) We find broad genome-wide correlation between association statistics for susceptibility to thirteen autoimmune and inflammatory diseases for which genome-wide association data were available (vitiligo, Vit; atopic dermatitis, AtD; allergy, All; asthma, Ast; ulcerative colitis, UC; inflammatory bowel disease, IBD; Crohn disease, CD; multiple sclerosis, MS; rheumatoid arthritis, RA; celiac disease, CeD; systemic lupus erythematosus, SLE; primary biliary cirrhosis, PBC; and systemic sclerosis, SSc). (B) This correlation is reflected in many loci harboring risk alleles to more than one of six diseases with available ImmunoChip data (lower triangle). Of these 236 pairs of associations, 99 are driven by the same underlying allele (upper triangle). (C) Risk alleles are mostly shared between two diseases (38 cases), with thirteen shared between three, and six between four diseases. (D) Twelve shared alleles have opposite effect directions, increasing risk of one disease and decreasing risk of another. This is most frequent between MS and IBD. (E) Combining cases and controls across diseases increases fine-mapping resolution for these shared associations. We assess resolution as the number of variants required to explain 95% of the posterior probability of association. This credible interval decreases by 58% when combining samples across diseases (red) compared to using only samples for one disease (orange). Associations that are not shared across diseases have similar credible interval distributions in individual diseases (blue).

While this shared heritability gives an overall impression of the relationship between diseases, it cannot identify specific genetic risk factors—and thus, genes and pathways—shared between diseases. To compare samples from different collections genotyped at different centers, it was important to minimize batch effects by ensuring all samples were profiled on the same platform. We therefore chose six autoimmune and inflammatory diseases with large numbers of samples genotyped on the ImmunoChip (*19*) (celiac disease, inflammatory bowel disease, multiple sclerosis, rheumatoid arthritis, systemic lupus erythematosus and type 1 diabetes; Fig. S1). This targeted array interrogates variants in 188 known risk loci to saturation, representing only 1.9% of the genome but capturing 38-86% of risk loci that have been identified in the six diseases (Fig. S3A). Using partitioned LD score regression, we confirmed that ImmunoChip regions account for 27.6% (MS) to 46.3% (CeD) of the estimated heritability for five of the six diseases for which GWAS data was available (Fig. S2B). After quality control, removal of population outliers, resolution of duplicate and related samples, and imputation to the 1,000 Genomes reference haplotypes, we analyzed a total of 104,302 SNPs in 188 non-MHC genomic regions for association with disease in 82,630 cases and 104,573 controls (Fig. S1; see Supplementary Text for additional details).

We first identified associations across the 188 loci in each disease independently by assembling cases and controls into homogeneous population strata and meta-analyzing across these groups. As multiple independent associations at a locus have been described in all diseases, we used stepwise logistic regression followed by fixed-effects meta-analysis to allow for such effects. We found 197 independent associations in 123 different ImmunoChip loci at genome-wide significance (*P* < 5 × 10^−8^), and 361 associations at 166 loci with suggestive association evidence (*P* < 10^−5^, Fig. S3B). Overall, we find some level of support for essentially all known genome-wide significant effects in the ImmunoChip regions.

We found substantial evidence for multiple independent associations within loci, with 7% (RA) to 30% (IBD) of loci exhibiting more than one independent effect (Fig. S3C). This included three instances of associations that have not been reported before (Fig. S4). In celiac disease, we found suggestive unconditioned associations at two loci: a variant intronic to *ANKS1A* on chromosome 6 (rs12206298; *P* = 4.1 × 10^−7^), and a variant intronic to *CTSH* on chromosome 15 (rs3784539; *P* = 1.3 × 10^−5^). After conditional association, both these associations passed the genome-wide significance threshold (*P* = 4.9 × 10^−8^ and 1.1 × 10^−8^ respectively). We found evidence of a second, independent effect in each locus (rs4713844, *P* = 9.9 × 10^−8^; and rs7181033, *P* = 8.7 × 10^−5^). Similarly, in IBD, we found that a suggestive association in the *CLEC16A* locus on chromosome 16 (rs7201325, *P* = 1.4 × 10^−7^) reached genome-wide significance after conditioning (*P* = 1.1 × 10^−10^), with evidence of a secondary, independent effect (rs55773334, *P* = 7.6 × 10^−5^). The presence of multiple masked independent effects highlights the need to look carefully at suggestive associations.

Having ensured we were capturing most of the known associations in ImmunoChip loci for each of the six diseases, we looked for shared effects across diseases, i.e. whether the same variant mediates risk to more than one disease. We found 224 overlapping conditionally independent associations at 98 loci (a lead variant associated to one disease at *P* < 10^−5^, and a lead variant for another disease *P* < 10^−4^; both lead variants being in LD *r*^*2*^ > 0.5 with at least one common SNP). Using joint likelihood mapping (JLIM), we found evidence of a shared effect in 93/224 (41.5%) such overlaps, involving a total of 56 conditionally independent shared effects spanning 53 unique loci (Figure 1B). Of these, 40 effects were shared between two diseases, ten between three, and six between four diseases (Fig. 1C). Unlike previous reports, which could not distinguish between shared and distinct associations with multiple diseases in a locus, these observations indicate that many mechanisms are shared between autoimmune and inflammatory diseases.

We found three loci where multiple conditionally independent associations for one disease were shared. In the *STAT4* locus, we found two independent effects each for RA and SLE were shared (Fig. S5A). In the *CD28*-*CTLA4* locus, one T1D risk association near *CD28* is shared with CeD, whereas another, an intronic variant in *CTLA4* is shared with RA (Fig. S5B). In the *TYK2* locus, one RA risk association is shared with SLE, IBD, and T1D; a second association, localizing to *ICAM3*, is shared with SLE alone (Fig. S5C). Cumulatively, these examples demonstrate that disease-associated alleles in the same locus can have different consequences, and that careful comparisons across diseases can distinguish each effect.

We next assessed if joint analysis across diseases could improve fine-mapping resolution. For each of the 56 shared associations, we assembled conditionally independent association data across all disease cohorts sharing that association, and combined them with fixed-effects, inverse variance-weighted meta-analysis. In a subset of loci, we saw an unexpected decrease in significance and increase in heterogeneity in the meta-analysis; we found these to be shared associations with opposite effects, where an allele increases risk for one disease and decreases it for the other (Fig. S6-S15). In six of these ten cases, variants with opposing effects were shared between MS and IBD. After inverting the association statistics to account for these effects, our meta-analysis resulted in higher significance for 123/134 (91.8%) associations across all 53 loci harboring a shared effect, demonstrating the potential to bolster association findings with our approach.

To establish if this increase in sample size provides a meaningful increase in fine-mapping resolution, we used FINEMAP (*20*) to calculate posterior inclusion probabilities for SNPs at each of the 56 shared effects, both in individual diseases and in the cross-disease meta-analysis. We then calculated 95% credible sets for each disease, both before and after cross-disease meta-analysis. We found a substantial decrease in the mean credible interval size, from 37.2 (s.d. 46.4) to 17.3 (s.d. 20.8), representing an improvement of 54% (Fig. 1E). We saw resolution improvement across the spectrum of initial association evidence, with the largest gains where an effect had relatively weak evidence of association in a disease: for associations below genome-wide significance in a single disease, our resolution increased from a mean of 51.8 SNPs to 18.6 SNPs after cross-disease meta-analysis; for associations already above genome-wide significance in a single disease, we saw improvement from a mean of 21.7 SNPs to 15.9 SNPs. This is exemplified by a shared association in the *C1orf106* locus on chromosome 1, where credible intervals of 28, 8, and 11 SNPs for CeD, IBD and MS respectively are reduced to eight variants in very tight linkage disequilibrium (minimum *r*^*2*^ = 0.976) on cross-disease meta-analysis (Fig. 2). In this case, there are genome-wide significant associations in each disease independently, but increasing sample size from symmetric equivalent 19,026 (CeD), 53,312 (IBD), 35,618 (MS) to a cross-disease meta-analysis 93,001 (symmetric equivalent) increases the resolution for both CeD and MS, identifying a core risk haplotype within *C1orf106*.

**Fig. 2:**
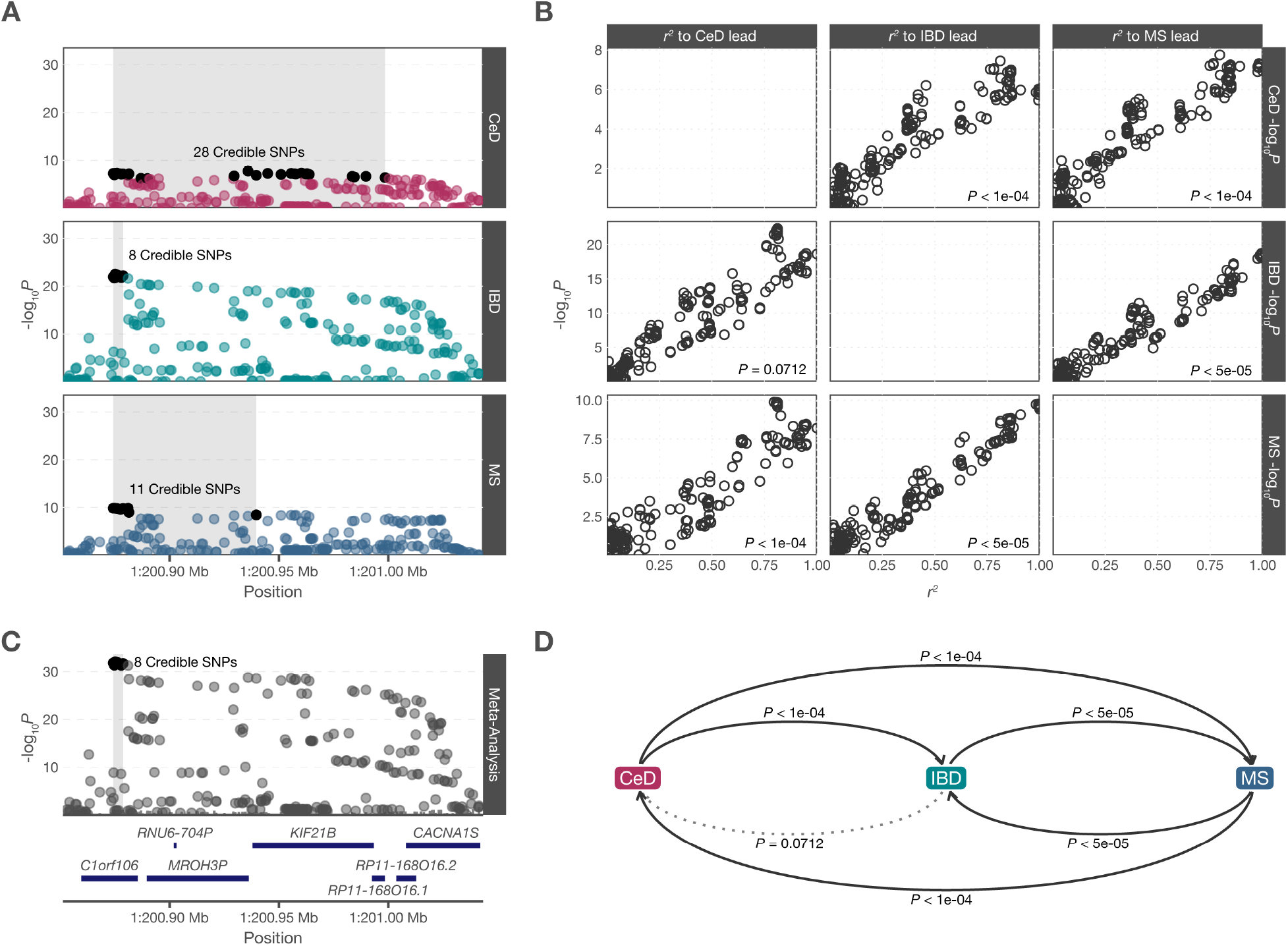
A shared effect on chromosome 1 can be fine-mapped to eight variants across celiac disease, inflammatory bowel disease, and multiple sclerosis. (A) Overlapping associations in celiac disease, inflammatory bowel disease and multiple sclerosis on chromosome 1, with 95% credible intervals varying both in number of variants and physical span. (B) For each pair of diseases, the strength of association (vertical axis) for the first trait decays in a linear fashion as a function of *r*^*2*^ to the lead SNP in the second trait, consistent with a shared causal variant. (C) Meta-analyzing across the three diseases gives in a stronger association signal, which can be fine-mapped to a narrow interval within *C1orf106*. (D) We find strong pairwise evidence that the association is shared between all three diseases; JLIM is asymmetric, so we run comparisons in both directions.

The ultimate promise of increasing fine-mapping resolution is to increase the interpretability of association signals. We and others have shown that disease risk associations are enriched in non-coding regions with gene regulatory potential (*4, 5, 21, 22*). We have used the JLIM approach to show that autoimmune disease associations are sometimes shared with expression quantitative trait locus (eQTL) signals, indicating the risk allele also influences gene expression. However, most associations are not shared with an eQTL, nor are they attributable to coding variants. To assess if this is due to limitations in fine-mapping resolution, we looked for shared associations between the 56 shared effects we discovered and *cis*-eQTLs for nearby genes in naïve T cells, monocytes and neutrophils in the BLUEPRINT dataset. We found 137 shared effects between each of 134 shared conditionally independent association signals in a single disease and eQTLs for nearby genes. We then looked for shared effects between the better-powered cross-disease meta-analysis data in each of the 53 loci, and can attribute 21 new disease/eQTL effects to the underlying diseases (Fig. 3; Table S3). Most of the implicated eQTLs are present in only one of the three cell types we interrogated, with T cells providing the largest number. We also exclude 11/137 disease/eQTL shared effects as no longer relevant because we do not find evidence of shared association between the cross-disease meta-analysis and eQTL data. Our gains primarily occur in cases where the cross-disease meta-analysis reduces the credible interval size (Fig. 3C), indicating that this gain of resolution drives these new observations.

**Fig. 3:**
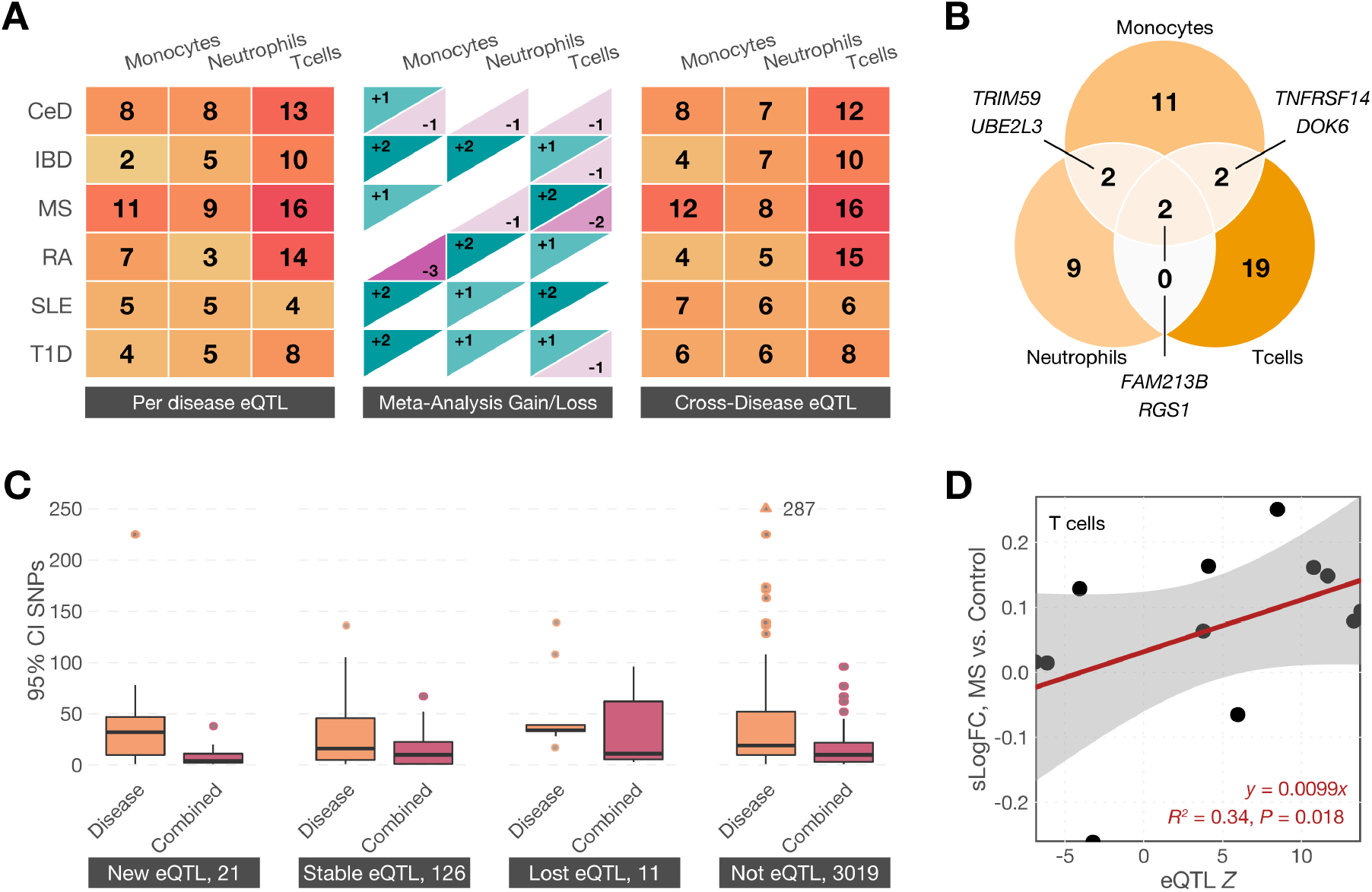
The increased resolution of fine-mapping shared associations across diseases allows identification of more disease-eQTL overlaps. (A) We looked for shared effects between disease associations and expression QTLs in loci harboring shared disease effects. When considering each disease separately, we find 139 significant disease—eQTL overlaps across monocytes, neutrophils and T cells from the BLUEPRINT consortium (left panel). When comparing eQTLs to cross-disease meta-analyses, we find new overlaps (blue, middle panel) and no longer find evidence for some eQTLs (red, middle panel), for a grand total of 157 disease-eQTL overlaps (13% net discovery increase, right panel). (B) Some of the shared eQTL effects can be detected in multiple tissues, but most are restricted to a single cell type, indicating substantial effect specificity. (C) We find new eQTL shared effects in loci where the cross-disease meta-analysis decreases the credible interval substantially, suggesting this resolution drives new discoveries. Disease associations where an eQTL is lost after meta-analysis also have smaller credible intervals, suggesting these may have been false positive findings due to lack of resolution in individual disease datasets. (D) The effects of risk-increasing shared alleles on gene expression is mirrored in expression differences between multiple sclerosis cases and controls. This suggests that risk states imparted due to small changes in gene expression persist during active disease, and provide validation that our eQTL discoveries are relevant to pathogenesis.

The direction of shared eQTL effects indicate whether we should expect increases or decreases in expression for those genes to increase disease risk. We reasoned that we might also see the same direction of effect between cases and controls, where the risk state is magnified. We therefore looked at single cell RNAseq data derived from T cells collected from a cohort of MS patients and healthy controls (*23*). After quality control, we were able to detect twelve genes that were targets of eQTLs shared with MS risk signals in our analysis. We found a significant pattern of correlation (*P* = 0.018): when a disease risk allele increased expression of a target gene, we saw higher expression in cases than in controls, and when it decreased expression we saw lower levels in cases than in controls (Fig. 3D). This suggests that shared associations do in fact drive risk-altering changes to gene regulation alter disease risk, and our results are uncovering pathogenic mechanisms.

The relative direction of the disease and eQTL associations can also suggest specific mechanistic hypotheses. This is exemplified by an association in the *RGS1* locus, shared between celiac disease and MS (Fig. 4). The cross-disease meta-analysis reduces the credible interval to 10 variants overlapping the promoter region of *RGS1*, which encodes a regulator of G-protein mediated signaling active in immune cell populations. We find a shared association between disease risk and *RGS1* expression in CD4 T cells, which is inverted so that lower expression correlates with higher disease risk. The lead credible interval variant overlaps a region of accessible chromatin within an active enhancer immediately upstream of the *RGS1* promoter. Further, this variant is annotated as a binding site for ZNF263 in the JASPAR database, and position-weight matrix analysis suggests the minor allele abrogates this binding site (*24*).

**Fig. 4:**
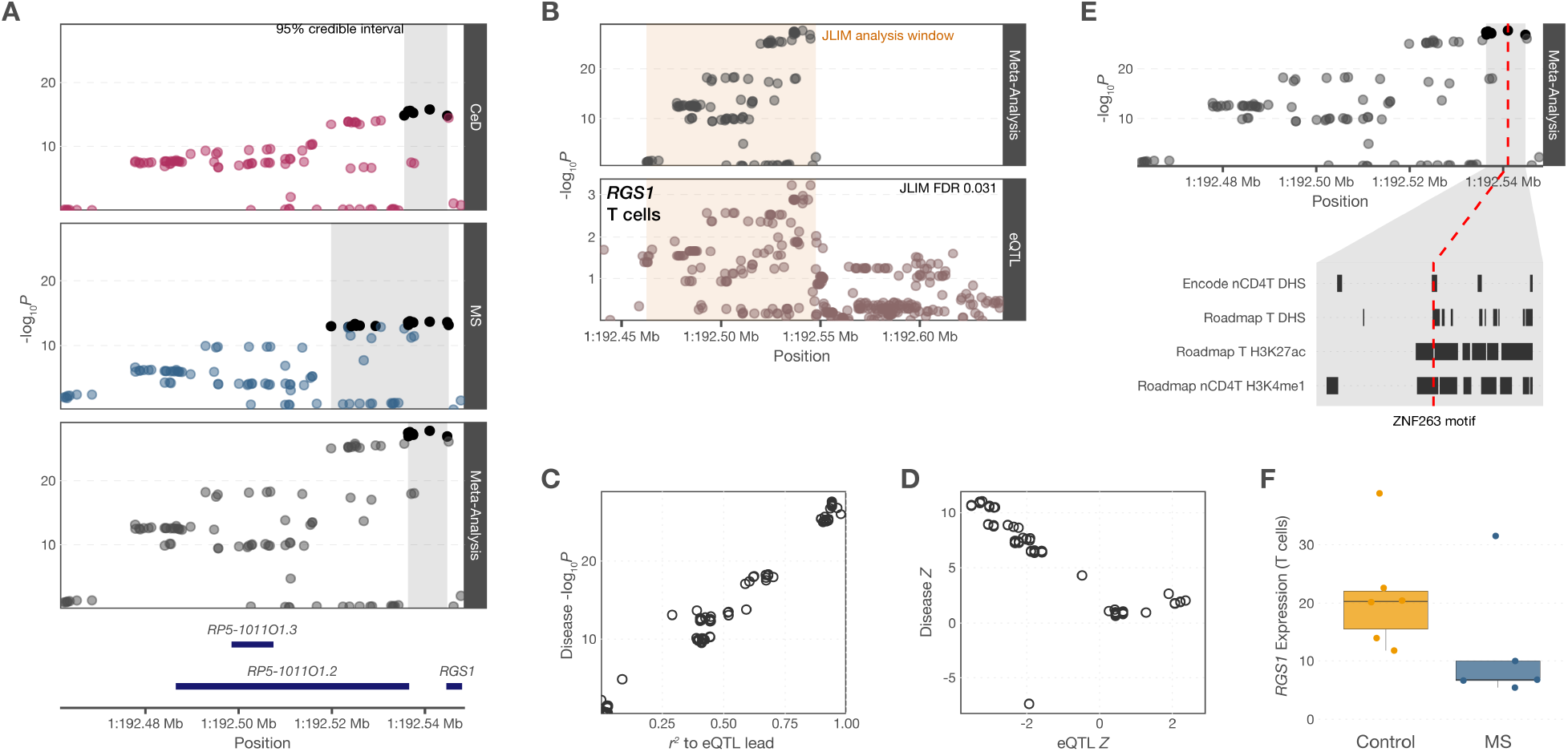
Jointly analyzing an association shared between multiple sclerosis and celiac disease improves fine-mapping resolution and identifies a shared eQTL for *RGS1*. (A) Overlapping associations for the two diseases are due to a shared effect (JLIM *P* = 5 × 10^−5^ for CeD as primary trait; *P* < 5 × 10^−5^ for MS as primary trait). Meta-analyzing across the two diseases increases the overall significance and a narrower credible interval (credible interval variants for each panel are in dark grey; the physical span of the credible interval is shaded grey). The credible interval focuses on the intergenic region proximal to *RGS1*. (B) This shared association is also shared with an eQTL for *RGS1* in naïve CD4 T cells (JLIM *P* = 0.015). (C) The lead disease-associated variant lies in a region of accessible chromatin in naïve CD4 T cells and total T cells. This is marked with H3K27ac in total T cells and with H3K4me1 in naïve CD4 T cells, suggesting this is an active, primed enhancer element. (D) The *RGS1* eQTL lead variant predicts the disease association *P* value, further indicating this is a shared effect. (E) Disease and eQTL association effects are negatively correlated, indicating that disease risk is associated with lower RGS1 expression. (F) *RGS1* is expressed at lower levels in T cells obtained from MS patients compared to healthy controls, confirming this risk effect direction.

We have quantified the shared heritability between autoimmune and inflammatory diseases, and demonstrated that we can leverage this to identify genetic variants that alter risk to multiple diseases. This significantly increases fine-mapping resolution, compared to the original genetic mapping studies: the number of effects where a single variant explains 95% of the posterior probability of association increases from 13 to 20 (a 54% increase); for 50% of the posterior probability, we see an increase from 35 to 54 (also 54%). Furthermore, we see an increase in the number of eQTLs, with evidence of sharing an effect with disease risk, from 139 to 154 (11%). Thus, in terms of identifying causal variants and functional interpretation, meta-analyzing across diseases meaningfully increases our ability to interpret genetic associations. This sets the stage for variant-to-function efforts to uncover key pathogenic mechanisms, as we provide high-value targets relevant to multiple diseases.

This approach can be applied to any set of traits sharing associations; we therefore suggest this is a fruitful avenue to maximize the interpretability of existing genetic studies of human complex traits, especially as shared mechanisms are applicable to multiple conditions. It is particularly valuable as sample collections, particularly of diseases that are difficult to diagnose or not especially common in the population, become depleted. Disease cohorts are often genotyped on different platforms, and the majority of common variants imputed. This can introduce a substantial bias, if cohorts of samples with different diseases have differential genome coverage. We have avoided this in our study by using a common platform, at the expense of not covering the entire genome. These technical hurdles will diminish as genotyping platforms coalesce around a standard set of variants, and as the community shifts to whole-genome sequencing rather than genotyping. We note that biological interpretation of genetic associations, shared or otherwise, is dependent on access to molecular and cellular phenotype studies such as eQTLs, which require profiling a wide array of tissues or cell types under diverse stimuli in order to identify the consequences of disease-associated variants. The BLUEPRINT dataset, which we used here, covers three very different blood cell types, but dozens more exist, in which the variants we have identified could act. This context specificity may be one reason we cannot always assign a cognate eQTL to each well-resolved association (*25*).

In terms of understanding the common mechanisms in autoimmunity, we and others have reported that many loci harbor associations to multiple autoimmune diseases. However, these approaches have relied on simple proximity of variants to infer that the underlying mechanisms must be shared. We have, for the first time, quantified the shared heritability between autoimmune and inflammatory diseases, and shown that a substantial proportion of shared loci harbor pleiotropic effects influencing risk to multiple diseases, which represent shared mechanisms. Many loci, however, harbor multiple independent effects, indicative of distinct mechanisms driving risk to different diseases; this is consistent either with the same underlying genes being influenced in different contexts to induce risk for different diseases, or with different genes which happen to be encoded near each other. Previous studies by us and others were not designed with this resolution, and could only identify loci harboring potentially different effects to multiple diseases.

Our results reveal complex patterns of shared heritability between autoimmune diseases. In particular, we find many opposite effects shared between IBD and MS, where the same allele increases risk for one disease but decreases risk for the other. This is reminiscent of the differential outcomes of anti-TNFα therapies, which are beneficial in IBD but exacerbate MS symptoms (*26*). Further, it suggests that some disease mechanisms may have an optimum state, and either hypermorphism or hypomorphism are deleterious. Overall, we see no evidence for a substantial component of risk shared across all six diseases, which would be indicative of a pan-autoimmunity mechanism. Our benchmarking suggests this is not due to a lack of power to detect shared effects (*27*), and our results strongly support independent effects in most loci. As our results argue against a single, shared autoimmune mechanism, they also dispute a single evolutionary origin for autoimmune and inflammatory diseases, which would have resulted in a set of risk alleles driving broad autoimmunity.

## Data Availability

All data analyzed are re-analyzed from publicly available sources. Detailed accessions are provided in Table S1.

## Acknowledgements

We thank the EAGLE eczema consortium for providing GWAS summary statistics. This research utilizes resources provided by the T1DGC, a collaborative clinical study sponsored by the National Institute of Diabetes and Digestive and Kidney Diseases (NIDDK), National Institute of Allergy and Infectious Diseases, National Human Genome Research Institute, National Institute of Child Health and Human Development, and JDRF and supported by U01 DK062418. We thank the RACI consortium for access to RA data and the International IBD Genetics Consortium for access to IBD data. De-identified data were provided from a total of 4617 samples (2563 SLE cases and 2054 population controls) in the Lupus Family Registry and Repository collection (*28*) at the Oklahoma Medical Research Foundation. The SLE Genentech samples were originally genotyped and analyzed as part of a large SLEGEN Consortium Immunochip study (*29*).

## Authors contributions

MRL, NC, CG, and MM curated and analyzed data;

DvH, CW, SW, IHJ, LP, IMSGC, SSR, RRG, PMG, CDL, and DAH provided data;

MRL and CC wrote and edited the manuscript, with input from all co-authors;

SC and SRS designed and implemented analytical methods;

CC conceptualized and oversaw the project.

## Data and materials availability

Dataset availability is documented in Table S1. Analysis source code is available from the Cotsapas lab GitHub page.

