## Supplementary methods for "Joint analysis reveals shared autoimmune disease associations and identifies common mechanisms"

**2 Supplementary Figures** **11**

**3 Supplementary Tables** **27**

**4 Supplementary Text: ImmunoChip Analysis** **31**
4.1 ImmunoChip Datasets and Quality Control . . . 31
*4.1.1 Manifest Resolution and Lift Over . . . 31*
*4.1.2 Stratum Identification . . . 32*
*4.1.3 Initial Missingness . . . 33*
*4.1.4 Sex Inconsistencies . . . 34*
*4.1.5 Population Outliers . . . 35*
*4.1.6 Initial Filtering for Hardy-Weinberg Equilibrium . . . 42*
*4.1.7 Filtering for Heterozygosity Outliers . . . 42*
*4.1.8 Strict Missingness . . . 46*
*4.1.9 Remove Duplicates . . . 48*
*4.1.10 Strict Filtering for Hardy-Weinberg Equilibrium . . . 49*
*4.1.11 QC Summary . . . 49*
4.2 Genotype Imputation . . . 50
*4.2.1 Pre-imputation filtering and strand consistency . . . 50*
*4.2.2 First Imputation . . . 51*
*4.2.3 Second Imputation . . . 54*
4.3 Association Analysis . . . 56
*4.3.1 Association Analysis . . . 56*
*4.3.2 Association Results . . . 57*
4.4 JLIM Simulations . . . 89
*4.4.1 Genotype Simulation . . . 89*
*4.4.2 Effect of Linkage Disequilibrium between Causal Variants . . . 91*

*4.4.3 Effect of Missing Genotypes and Imputation . . . . . 93*
*4.4.4 Effect of Multiple Causal Variants at a Single Locus . . . . . 95*
4.5 Identification of Colocalized Traits . . . . . 100
*4.5.1 Identification of Candidate Trait Pairs . . . . . 100*
*4.5.2 JLIM Analysis . . . . . 100*
4.6 Cross-Disease Meta-Analysis and Fine Mapping . . . . . 101
*4.6.1 Meta Analysis Across Colocalized Traits . . . . . 101*
*4.6.2 Fine-Mapping . . . . . 115*

**References 127**

### Section 1

### Materials & Methods

#### 1.1 Shared Heritability Analysis

We downloaded complete summary statistics for all autoimmune and inflammatory disease GWAS available in the NHGRI-EBI GWAS catalog (*30*). We focused on European ancestry studies with at least 2,000 subjects for which signed summary statistics were available. Where multiple studies were available for a given trait, we chose the study with the largest cohort size. By applying these filters, we obtained GWAS statistics for atopic dermatitis (AtD) (*31*), allergic traits (All) (*32*), asthma (Ast) (*33*), celiac disease (CeD) (*34*), eosinophilic granulomatosis with polyangiitis (EGPA) (*35*), selective IgA deficiency (sIgAD) (*36*), inflammatory bowel disease (IBD) (*3*), latent autoimmune diabetes in adults (LADA) (*37*), primary biliary cirrhosis (PBC) (*38*), primary sclerosing cholangitis (PSC) (*39*), psoriatic arthritis (PsA) (*40*), systemic lupus erythematosus (SLE) (*41*), systemic sclerosis (SSc) (*42*), and vitiligo (Vit) (*43*). IBD summary statistics also included results for Crohn's disease (CD) and ulcerative colitis (UC). We downloaded summary statistics for psoriasis (Ps) (*44*) from dbGaP and summary statistics for rheumatoid arthritis (RA) (*45*) from GRASP. We obtained multiple sclerosis (MS) summary statistics from the International MS Genetics Consortium (*2*). Sources and accession numbers for included studies are documented in Table S1.

We first removed indels and single nucleotide polymorphisms (SNPs) inconsistent with the 1,000 Genomes Project (Phase 3) reference panel (*46*). We next filtered for strand-unambiguous biallelic SNPs with minor allele frequency (MAF) > 0.01 in the 1,000 Genomes European (EUR) reference subjects. Following Bulik-Sullivan et al. (*15*), we removed variants with INFO < 0.9 where this information was available. As INFO scores were not available for most datasets, we uniformly filtered on SNPs present in the HapMap 3 (*47*) reference panel. Where differing effective sample sizes were provided for each variant, we removed SNPs genotyped in fewer than two-thirds of the 90th percentile population size.

After quality control, we used linkage disequilibrium (LD) score regression (*14*) to estimate heritability ($h_g^2$) for each trait from summary statistics, using the 1,000 Genomes EUR individuals as reference. We excluded traits with heritability $Z$-scores < 4 from further analysis. We next used LD score regression to calculate correlations ($r_g$) among all pairs of the remaining traits.

#### 1.2 ImmunoChip Datasets

We obtained raw ImmunoChip genotypes for six autoimmune and inflammatory diseases (Figure S1; Table S1), including CeD (*48*), IBD (*49*), MS (*50*), RA (*51*), SLE (*29*, *52*) and type 1 diabetes (T1D) (*53*). Each of the participating disease consortia provided data, including two separate SLE consortia (OMRF and Genentech). For the CeD and SLE datasets, we used GenomeStudio to call genotypes from intensity files.

After resolving conflicting SNP nomenclature and allelic encoding across datasets, we lifted these over from GRCh36 (hg18) to GRCh37 (hg19). We excluded SNPs that could not be mapped unambiguously to the newer assembly.

All datasets consisted of multiple strata, typically divided by country of origin (Table S2). We therefore divided datasets into country-level strata and performed quality control independently within each stratum. As one of the T1D datasets consisted of affected sibling pairs, we processed this separately.

#### 1.3 Genotype Quality Control

Quality control and association analysis are discussed in detail in the Supplementary Text (Section 4) and here in brief. We used `PLINK` (*54*) to perform initial quality control. Within each stratum, we first removed individuals missing >10% of genotypes, and SNPs that were missing in >5% of individuals. We then assessed the remaining samples for sex inconsistencies. Where X chromosome genotypes were available, we calculated X chromosome homozygosity for each individual. We then used `Mclust` (*55*) to divide samples into male and female clusters assuming a Gaussian mixture model with two components. Inferred sex was used where these were not specified in the original datasets. Individuals were removed if their recorded sex differed from the model-inferred sex.

We focused our analysis on individuals of European descent. To identify population outliers, we merged our genotype data with reference data from the 1,000 Genomes Project (*46*) and performed principal component analysis. Plotting samples by their first two principal components (*56*), we removed samples in an iterative fashion, first removing individuals closer to EAS or AFR than to EUR. We then repeated principal component analysis and removed samples closer to SAS than EUR. At each step, samples that did not correspond to an identifiable reference population were removed empirically.

After removing population outliers, we removed SNPs exhibiting deviation from Hardy-Weinberg equilibrium expectation ($P < 10^{-8}$). We next identified and removed subjects with extreme homozygosity. We used `PLINK` to calculate inbreeding coefficients (F) for each individual. Within each stratum, we removed individuals with F > 2.5 s.d. from the stratum mean. We next applied a second, more stringent filter for missing values, removing individuals with >1% missing data and SNPs missing in >1% of individuals.

We then identified close relatives ($\hat{\pi} > 0.185$) and duplicates ($\hat{\pi} > 0.90$) within each disease dataset. Duplicates were removed from further analysis. Relatives were excluded from a second Hardy-Weinberg equilibrium assessment, where we filtered SNPs that violated Hardy-Weinberg equilibrium at $P < 10^{-5}$. We included relatives to provide additional chromosomes for phasing and imputation; we removed these before association testing. After quality control, we excluded strata with fewer than 150 remaining cases or controls.

A total of 168,928 subjects were available for analysis after quality control. After identifying and removing control samples that were shared among studies, we were left with 129,058 unique individuals. The numbers of subjects supplied and analyzed are indicated in Figure S1. More detail on the numbers of subjects per stratum are given in Table S2. A detailed accounting of all subjects (Data S1) and SNPs (Data S2) can also be found in the Supplementary Data.

#### 1.4 Imputation and Association Analysis

Before imputation, we removed indels, rare SNPs (MAF < 0.05) and SNPs that were missing differentially between cases and controls ($P < 10^{-5}$). We then used `SHAPEIT2` (*57*) to remove SNPs that were inconsistent with the 1,000 Genomes (Phase 3) reference haplotypes and to phase remaining SNPs. We used `IMPUTE2` (*58*) to impute reference SNPs that were not genotyped, and to fill in sporadically missing genotypes. A subset of genotyped SNPs could not be reliably imputed from the reference haplotypes (`concord_type0` < 0.75 despite `info_type0` > 0.8). As these were either mis-mapped or unreliably genotyped, we excluded these SNPs and performed a second round of imputation as above.

We removed imputed SNPs if they did not have two alleles present, if they were imputed with `INFO` < 0.75, or if they had MAF < 0.05. We also removed variants that violated Hardy-Weinberg equilibrium at $P < 0.001$, and those that exhibited differential missingness between cases and controls at $P < 0.01$.

We used SNPTEST (*59*) to perform logistic regression of imputed genotype dosages against phenotype in each stratum, incorporating the first two principal components as covariates into an additive model (see Supplementary Text, Section 4.3.1). We then combined association statistics into a fixed-effects, inverse variance-weighted meta-analysis (*60*) for each disease. The extended MHC (6:28-34 Mb, GRCh37 coordinates) was excluded from analysis.

To allow for multiple independent effects at a given locus, we used iterative stepwise conditional logistic regression. For each iteration after the first, we repeated logistic regression in each stratum, this time conditioning on all previously identified meta-analysis lead SNPs with $P < 0.0001$. Results were again combined in a fixed-effects meta-analysis. We restricted our search for lead variants to SNPs present in all strata, with $I^2 < 50$ and $r^2 < 0.9$ to all previous lead SNPs. Where such a lead SNP could be identified, we added this to the list of conditioning variants and proceeded with another round of association testing. We continued conditioning until we detected three independent signals or no variants with $P < 0.0001$ remained.

Our iterative conditioning approach produced a set of independent associations for each trait at each ImmunoChip locus. To identify conditionally independent association signals at each locus, we iterated over the set of lead variants, this time conditioning on the all-but-one variant. For this analysis, we again required SNPs to be present in all strata, with $I^2 < 50$.

##### 1.4.1 Identification of Shared Genetic Effects

We used Joint Likelihood Mapping (JLIM) (*27*) to identify genetic effects that are shared across multiple diseases at each ImmunoChip locus. The method relies on permutation of genotype-level data, so we restricted our analysis to diseases with data available for large numbers of samples. We wished to analyze trait pairs exhibiting at least moderate strength of association; we therefore identified pairs of traits at each locus with lead variants significant at $P_1 < 0.00001$ and $P_2 < 0.0001$. To ensure a moderate degree of linkage disequilibrium between assessed traits, we identified the set of variants with $r^2 \geq 0.5$ to each lead variant; trait pairs were assessed for a common underlying genetic effect where these sets shared at least one variant. To this initial set of candidates, we added additional trait pairs that appeared similar based on their Manhattan plots.

For each analyzed trait pair, we identified and removed shared controls before `JLIM` analysis. We then applied `JLIM` in both directions, using each trait alternately as the primary and secondary trait. We set the analysis window to be the maximal coordinates of the union of the $r^2 \geq 0.5$ windows; resolution was set to the default $r^2 = 0.8$. Linkage disequilibrium in the primary trait was estimated from 1,000 Genomes reference data; for the secondary trait, we estimated this directly from best-guess genotypes.

We estimated JLIM significance by permutation. For each permutation, we shuffled phenotype labels independently in each disease stratum and repeated logistic regression and meta-analysis as above. A minimum of 10,000 permutations were performed for each trait pair.

To identify clusters of traits sharing a common genetic effect, we analyzed pairwise `JLIM` results as graphs. Edges were defined between traits where `JLIM` was significant at $P < 0.05$. Maximal connected undirected subgraphs were then identified at each locus. For subgraphs of size greater than two, we repeated `JLIM` for each ordered pair of traits in the subgraph, this time using a common analysis window defined by the union of the $r^2 \geq 0.5$ windows for all traits in the subgraph.

#### 1.5 Fine-Mapping of Susceptibility Loci

For each cluster of traits sharing a common genetic effect, we combined data by meta-analysis. Duplicate samples were identified and removed from each cluster. We then repeated logistic regression and meta-analysis as described above. We used the $I^2$ statistic to assess variants for heterogeneity. Within each disease, variants were excluded if $I^2 \geq 50$, or if they were present in fewer than half of the constituent strata. To be included in fine-mapping analysis, variants were required to have survived filtering in all diseases of a given cluster.

Trait pairs with opposing effect directions were identified by linear regression. For each pair, we regressed SNP $Z$ scores for the first trait against corresponding $Z$ scores for the second trait. We considered trait pairs to have opposing effect directions when their slope term was negative and statistically significant. For such trait pairs, we reversed the direction of the effect for one trait and repeated meta-analysis. Opposing trait pairs were confirmed by comparing heterogeneity statistics before and after reversal.

For each shared effect cluster, we used `FINEMAP` (*20*) to estimate posterior inclusion probabilities for each variant within our shared effect clusters. SNP correlation matrices were calculated from genotypes for each trait. We used shotgun stochastic search, assuming a single causal variant at each locus. We quantified fine-mapping improvement by comparing the number of SNPs in 95% credible intervals for individual disease traits, and for meta-analyzed clusters.

#### 1.6 Expression Quantitative Trait Locus (eQTL) Data

We obtained and quantitated raw RNA-sequencing reads from three human immune cell types from the BLUEPRINT consortium (*61*): neutrophils (CD66b+ CD16+; 196 individuals), monocytes (CD14+ CD16-; 193 individuals), and naïve CD4 T cells (CD4+ CD45RA+; 169 individuals). Subjects in this study were ascertained to be free of disease and were representative of the United Kingdom population. We used the GTEx Analysis V8 pipeline to align FASTQ files, filter for quality control and quantitate gene expression. Briefly, we used `STAR` v2.5.3a to align reads to GRCh38. We quantitated expression to the gene level with `RNA-SeQC` v1.1.9, using the GENCODE 26 gene model. We included genes with expression values >0.1 TPM and ≥6 reads in at least 20% of samples; we then normalized counts using the trimmed mean of M-values (TMM) method implemented in `edgeR` (*62*). We then normalized expression across samples using an inverse normal transformation. We retained all samples for analysis as none had fewer than the minimum 10 million reads.

We obtained genotype data for all individuals with available gene expression data; a total of 7,008,524 variants were available. Whole-genome sequencing, alignment, variant calling and quality control were performed previously (*61*). All SNPs were biallelic. We removed indels and SNPs with MAF < 0.05 or that violated Hardy-Weinberg equilibrium at $P < 0.00001$. There were no heterozygosity outliers, defined as samples with heterozygosity > 5 standard deviations from the sample mean. Similarly, there were no cryptic relatives ($\hat{\pi} > 0.1875$) or population outliers (>4 standard deviations in the first four PCs). A total of 4,853,096 variants were available for analysis in 197 subjects.

#### 1.7 Identification of Shared Susceptibility-eQTL Loci

We used `JLIM` to identify shared eQTL–disease susceptibility loci. Disease susceptibility summary statistics were lifted over to GRCh38 coordinates. For each shared effect, we assessed all genes with transcription start site within 1 Mb of any susceptibility lead SNP. We regressed normalized expression values for these genes against genotype in a linear model, assuming an additive model of inheritance. We used covariates to adjust for age, sex the first 5 principal components and 30 PEER factors (*63*). The same covariates were used to generate permutation data for `JLIM`.

To allow for multiple independent eQTLs within a given locus, we performed conditional cis-eQTL analyses. For eQTL with $P < 0.001$, we repeated linear regression modelling, this time conditioning on the lead SNP from the first model. We continued adding lead variants to our model until either (a) the lead variant $P \geq 0.001$ or (b) three conditioning SNPs had been included. To identify conditionally independent eQTL signals, we again iterated on the set of lead variants, this time conditioning on the all-but-one variant.

After identifying conditionally independent eQTL signals for each gene, we used JLIM to assess for a common underlying genetic effect between disease susceptibility loci and eQTLs. We lifted summary statistics for susceptibility loci over to GRCh38 and used these as primary traits. Expression QTLs were used as secondary traits. For each primary trait, the JLIM analysis window was chosen to be the union of all SNPs ±100 kb from the lead SNP. We estimated significance by permuting eQTL expression values 100,000 times for each trait. Within a given cluster of disease associations, we used the Benjamini-Hochberg procedure to correct *P*-values for the number of genes and cell types assessed.

#### 1.8 Single Cell RNA-seq Analysis

##### 1.8.1 Sample Quality control

We obtained raw sequencing data from a previously-published single cell RNA-seq (scRNA-seq) study of multiple sclerosis (*23*). Sample collection and data preparation are described in detail in the original publication. Briefly, peripheral blood mononuclear cells (PBMCs) and cerebrospinal fluid (CSF) cells were obtained from 6 healthy donors and 5 new-onset multiple sclerosis patients. For each donor, single cell suspensions were prepared for analysis using the 10x Genomics platform. We used unique molecular identifier (UMI) count matrices as described (*23*). We filtered extreme outliers by excluding droplets with (a) <1000 UMI counts or <500 unique genes detected, or (b) >15,000 UMI counts or >5,000 genes detected. To exclude low-quality cells and potential doublets from our analysis, we examined the distributions of UMI counts and number of detected genes per cell. As distributions of these parameters varied across emulsions, we quantile-normalized $\log_{10}$-transformed UMI counts and $\log_{10}$-transformed number of detected genes per cell. Using quantile-normalized values and the percentage of counts mapping to mitochondrial genes, we excluded low-quality cells with <2,000 UMI counts, <900 genes detected, or >12.5% counts mapping to mitochondrial genes. We also excluded doublets with >8,000 UMI counts or <2000 genes detected.

##### 1.8.2 Dimensionality Reduction and Clustering

For cells passing quality control, we normalized UMI counts using a count per million approach, dividing each count by the total number of counts per cell. We then multiplied normalized counts by 10,000 and added a pseudo count of 1 before log-transformation. We then applied a variance-stabilizing transformation (VST) to account for variation in gene expression levels across the dataset, and used genes with stabilized variance >1 and stabilized mean expression $>10^{-3}$ as input for principal component analysis (PCA). Genes mapping to the T cell receptor (TCR), the B cell receptor (BCR) and the Y chromosome were excluded from PCA. We computed the first 50 principal components (PCs) using a partial singular value decomposition method, based on the implicitly restarted Lanczos bidiagonalization algorithm (IRLBA), as implemented in the `Seurat` package (*64*).

To correct for systematic differences across samples we applied `Harmony` integration (*65*) to the first 50 PC loadings. We then retained the first 30 harmony-corrected PCs, and used PC loadings as input for visualization using UMAP (minimum distance = 0.5, spread = 10), and clustering by applying the Louvain algorithm to a shared nearest neighbors (SNN) graph (resolution = 0.01), as implemented in `Seurat`. This low-resolution clustering separated T and NK cells from B cells and monocytes. We then selected T and NK cells and re-applied the same pipeline to the raw UMI counts to obtain a dedicated UMAP visualization and clusters of T cells (SNN $k$ = 20 and Louvain resolution = 0.5), enabling us to distinguish between different T cell sub-populations. Using normalized log-transformed UMI counts, we computed the area under a receiver operating curve (auROC) to define diferentially expressed genes between each cluster pairs. Manual inspection of gene markers enabled us to define several sub-populations (by order of abundance): central memory CD4 T cells (cluster 0: *CD4*, *CXCR5*, *LTB*, *KLRB1*), naïve CD4 T cells (cluster 1: *TCF7*, *CCR7*, *LEF1*, *CD4*, *STAB1*, *TSHZ2*, *NPM1*, *SELL*), central memory CD8 T cells 1 (cluster 2: *CCL5*, *CD8A*, *GZMA*, *NKG7*, *GLNY*, *CD8A*), naïve CD8 T cells (cluster 3: *TCF7*, *NELL2*, *SELL*, *CCR7*, *LEF1*, *CD8A*, *CD8B*), effector CD8 T cells 1 (cluster 4: *CCL4*, *CCL5*, *GZMA*, *CST7*, *PRF1*, *CD8A*, *CD8B*), natural killer cells (cluster 5: *KLRG1*, *NKG7*, *PRF1*, *KLRB1*, *GZMK*), regulatory T cells (cluster 6: *FOXP3*, *IL10RA*, *TIGIT*, *CD4*), gamma-delta T cells (cluster 7: *TRDC*, *KLRB1*, *GLNY*, *KLRC1*, *CCL5*, *GZMA*, *PRF1*), T follicular helper CD4 T cells (cluster 8: *FAU*, *FTH1*, *VIM*, *CD4*), megakaryocytes (cluster 9: *NRGN*, *PPBP*, *TUBB1*, *SPARC*), type I interferon activated CD4 T cells (cluster 10: *MX1*, *ISG15*, *IRF7*, *XAF1*, *IFI6*), central memory CD8 T cells 2 (cluster 11: *ZNF683*, *CD7*, *KLRC3*, *LEF1*, *CD8A*, *CD8B*), central memory CD8 T cells 3 (cluster 12: *TCF7*, *CD27*, *GZMK*, *CD8B*).

##### 1.8.3 Pseudo-Bulk Analysis

We used cluster assignments to sum UMI counts across cell types, disease status, tissue and donor. For each cell type in the blood compartment (PBMCs), we used a negative binomial distribution with a local fit, as implemented in `DESeq2` (*66*) to model gene expression and test differences between cases and controls, while controlling for sex as a covariate. We used shrinkage to account for $\log_2$-fold change inflation on genes with low counts and used shrunken $\log_2$-fold change for subsequent analyses. We focused on cluster 0 for validation of T cell eQTL predictions as this cluster was most abundant.
