## supplementary figures for "Joint analysis reveals shared autoimmune disease associations and identifies common mechanisms"

### Section 2

### Supplementary Figures

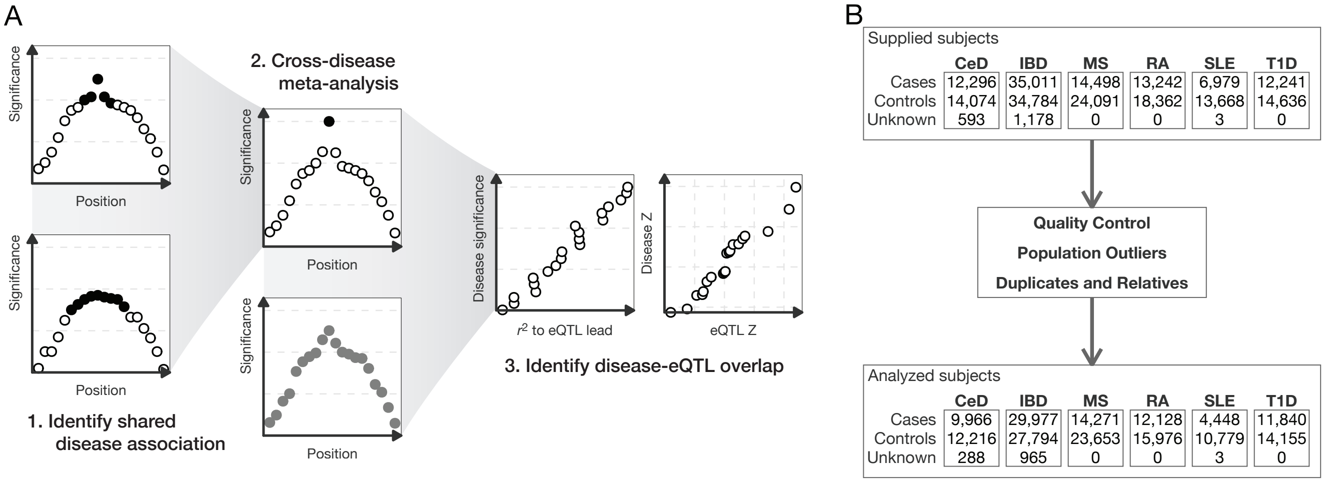


**Fig. S1:** Methods overview. (A) Schematic overview of cross-disease fine-mapping. We used joint likelihood mapping (JLIM) to identify susceptibility loci that are likely to share a common underlying causal variant across multiple autoimmune diseases (left panels). At such loci, we performed cross-disease meta-analysis, combining data for co-localized diseases into a fixed-effects model (middle panel, top). We then performed statistical fine-mapping; SNPs contained within the 95% credible interval are shown as filled black circles in this schematic. In general, cross-disease fine-mapping produced smaller credible intervals, in this schematic represented as a single causal variant. We next used `JLIM` to assess each susceptibility locus for overlap with cis-eQTLs in the BLUEPRINT dataset of naïve CD4 T cells, monocytes and neutrophils. In this schematic, the meta-analysis signal overlaps with an eQTL signal (lower middle panel). At such overlaps, we expect the disease susceptibility signal to decay as a linear function of correlation to the eQTL lead variant. We also expect a linear correlation between corresponding effect sizes for susceptibility and eQTL gene expression (right panels). (B) Overview of the number of subjects assessed and which passed quality control.

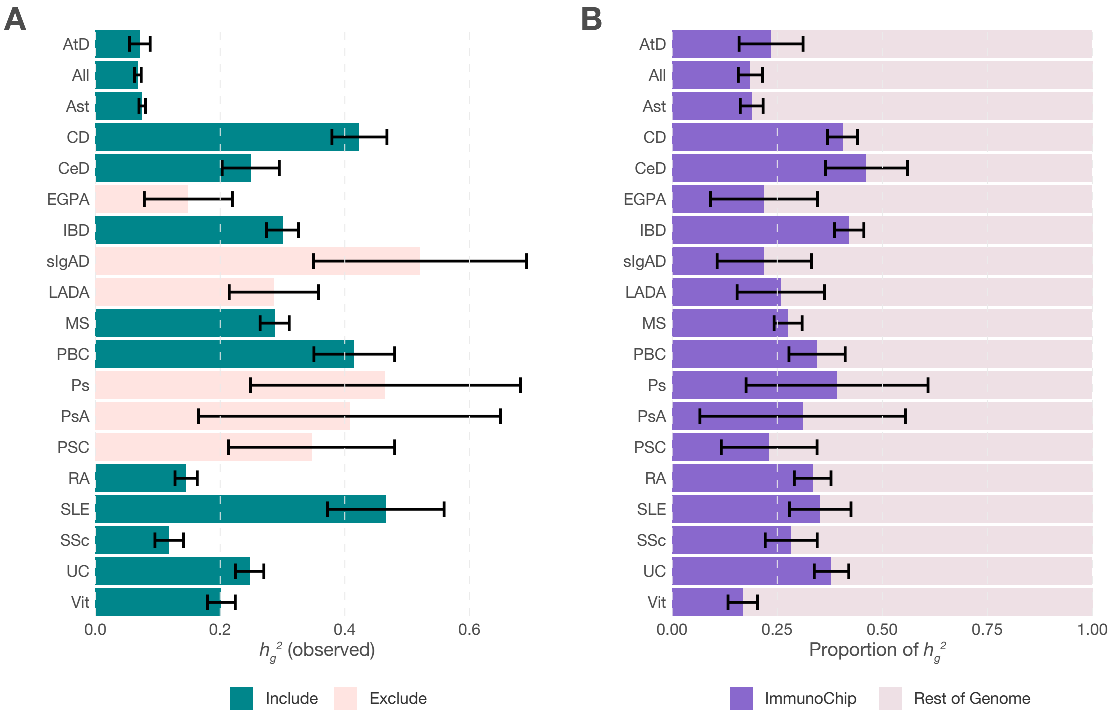


**Fig. S2:** Heritability of immune-mediated disorders is enriched in ImmunoChip regions. We used LD score regression to estimate heritabilities ($h_g^2$) for 19 immune-mediated disorders for which GWAS summary statistics were available. We excluded traits with heritability $Z$-scores < 4 (indicated in pink) from further analysis. (A) Heritability estimates for the remaining 13 traits (observed scale) are highly variable, ranging from 0.068 (All) to 0.47 (SLE). While heritability is sensitive to population and method of estimation, we see that several estimates are smaller than expected, reflecting the influence of genomic control correction used in the original association studies. As this downward bias affects both numerator and denominator equally (*15*), it does not influence genetic correlation analysis. (B) Using partitioned LD score regression (*6*) to measure the proportion of heritability that can be attributed to ImmunoChip regions (~2% of the genome), we see broad patterns of enrichment, ranging from 16.8% (Vit) to 46.3% (CeD).

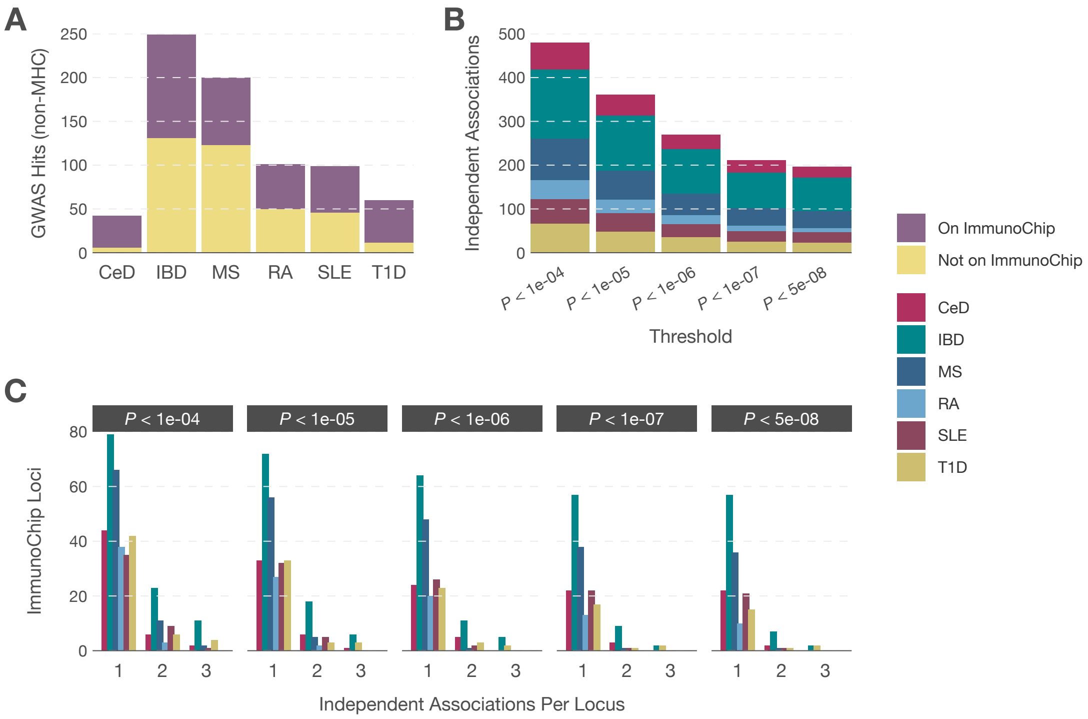


**Fig. S3:** ImmunoChip covers a significant fraction of GWAS loci for six autoimmune diseases. (A) Between 38.5% (MS) and 85.7% (CeD) of previously identified, non-MHC GWAS susceptibility loci for CeD, IBD, MS, RA, SLE and T1D fall within high-density ImmunoChip regions. (B) The total number of conditionally independent associations declines as a function of *P*-value threshold, with 495 independent effects identified at $P < 0.0001$ and 202 independent effects identified at $P < 5 \times 10^{-8}$. At all thresholds, the largest number of effects are identified for IBD, the largest dataset. (C) The majority of associated loci exhibit a single genetic effect at all thresholds. At $P < 5 \times 10^{-8}$, up to 13.6% of associated loci in IBD exhibit more than one effect; at the lowest threshold, multiple effects are seen at between 7.3% (RA) and 30.1% (IBD) of associated loci.

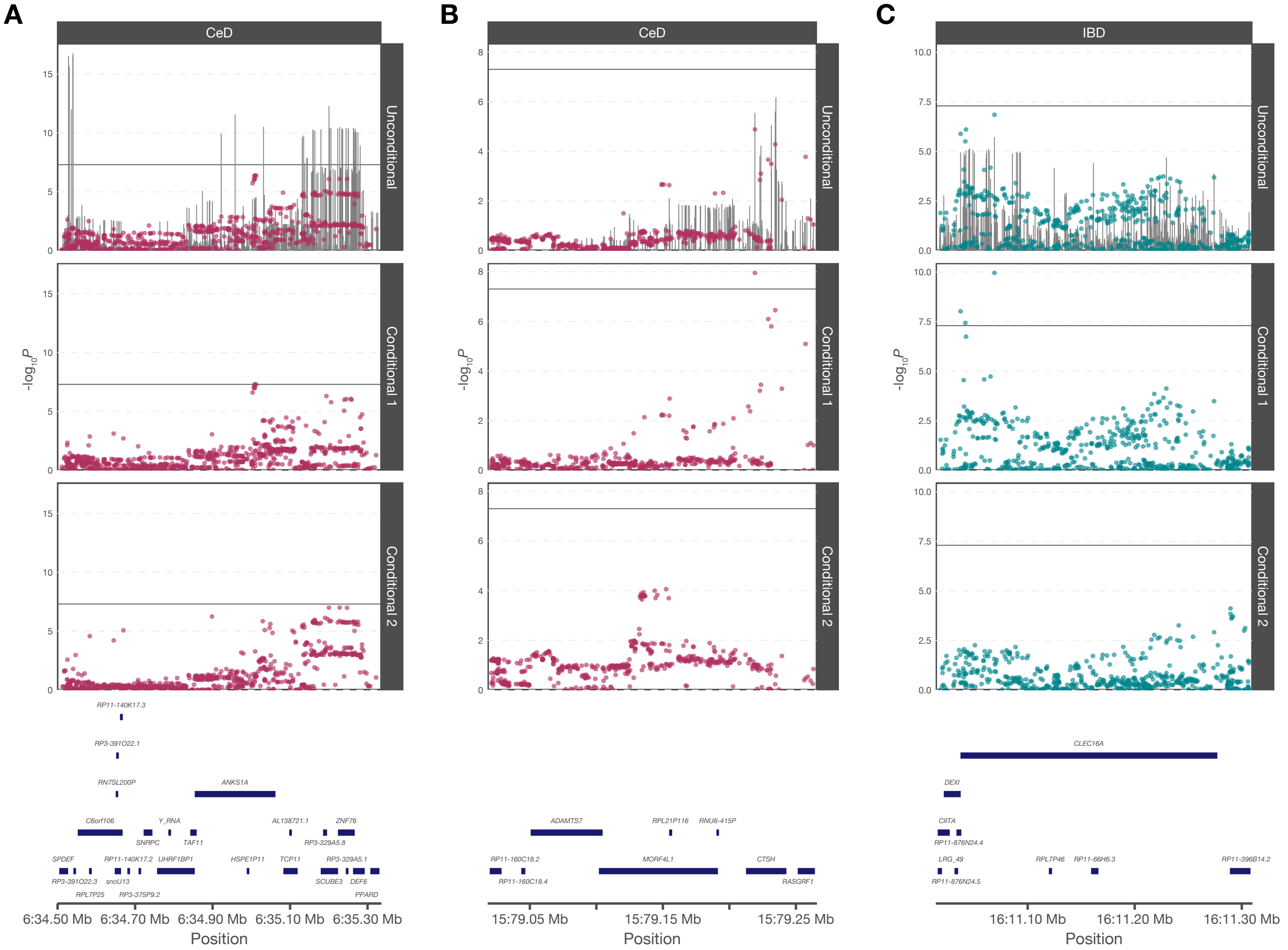


**Fig. S4:** Previously unreported ImmunoChip associations. Using conditional logistic regression to allow for multiple associated variants at a single locus, we identified three genome-wide significant associations (two for CeD, one for IBD) that were not reported in the respective publications. (A) Unconditional testing at the *ANKS1A* locus on chromosome 6 produced evidence of association that did not reach genome-wide significance in our analysis. Conditional testing produced genome-wide significance for a single variant within *ANKS1A* (rs12206298; $P = 4.9 \times 10^{-8}$), and suggestive evidence for a second effect (rs4713844; $P = 9.9 \times 10^{-8}$) in the region. Published summary statistics (lines, top panel) were also significant. The region may have been excluded from the initial publication as it is within the extended MHC. (B) Unconditional testing at the *ADAMTS7*—*MORF4L1*—*CTSH* locus on chromosome 15 in CeD did not reach genome-wide significance. Conditional testing revealed genome-wide significance for a variant in *CTSH* (rs3784539; $P = 1.1 \times 10^{-8}$) and suggestive evidence for a second effect in *MORF4L1* (rs7181033, $P = 8.7 \times 10^{-5}$). (C) Unconditional testing at the *CLEC16A* locus on chromosome 16 in IBD did not reach genome-wide significance. Conditional testing produced evidence for a single effect in *CLEC16A* (rs7201325, $P = 1.1 \times 10^{-10}$) and modest evidence for a second effect near *RP11-396B14.2* (rs55773334, $P = 7.6 \times 10^{-5}$).

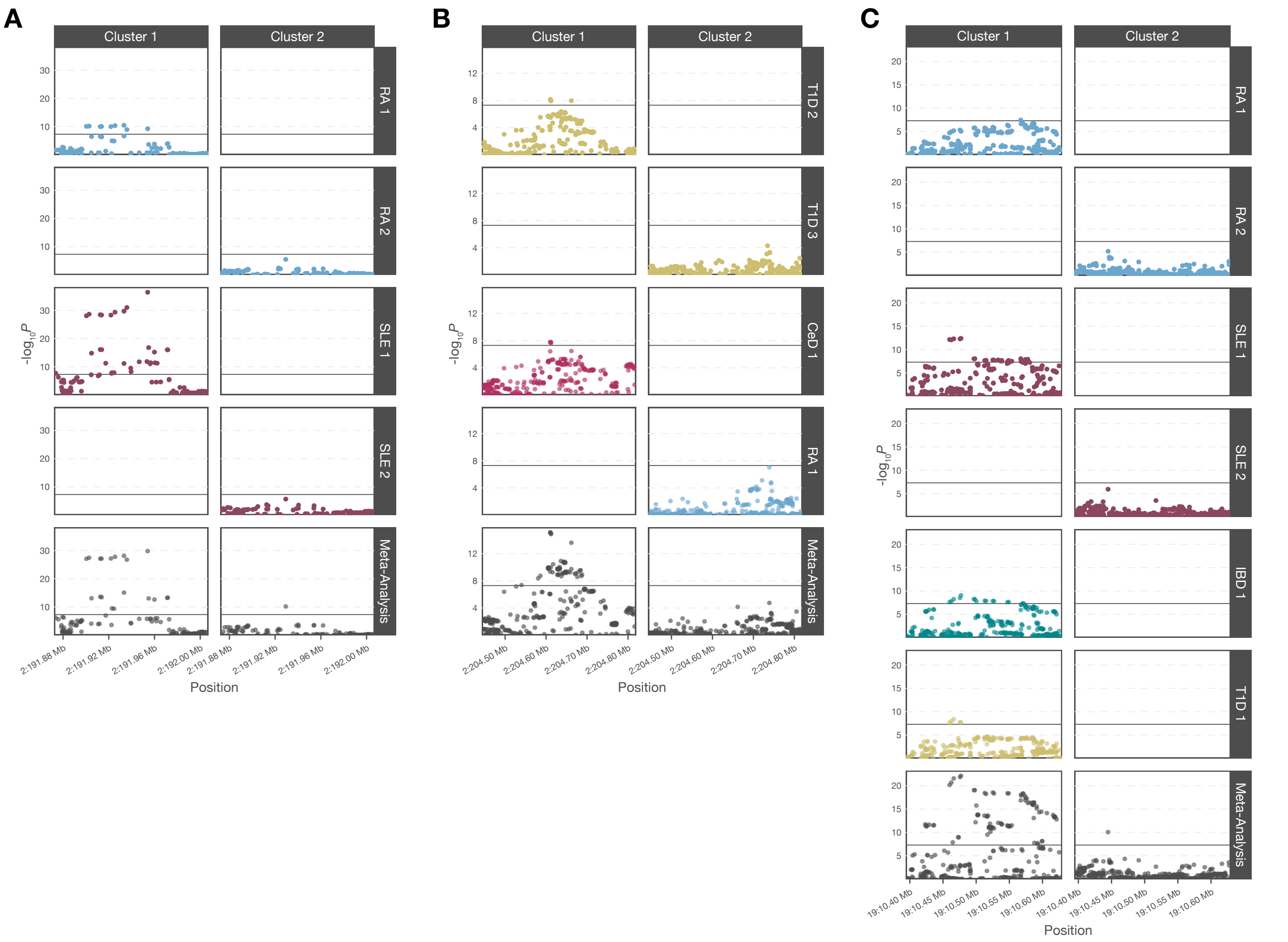


**Fig. S5:** Distinct conditional associations are shared with distinct sets of diseases. At three loci, multiple conditionally independent associations for a single disease are shared. (A) At the *STAT4* locus, two independent effects are each shared between RA and SLE. (B) At the *CD28-CTLA4* locus, the second independent association in T1D near *CD28* is shared with CeD and the third independent association within *CTLA4* is shared with RA. (C) At the *TYK2* locus, the first independent association of RA is shared with SLE, IBD and T1D, while the second association in RA (near *ICAM3*) is shared with SLE.

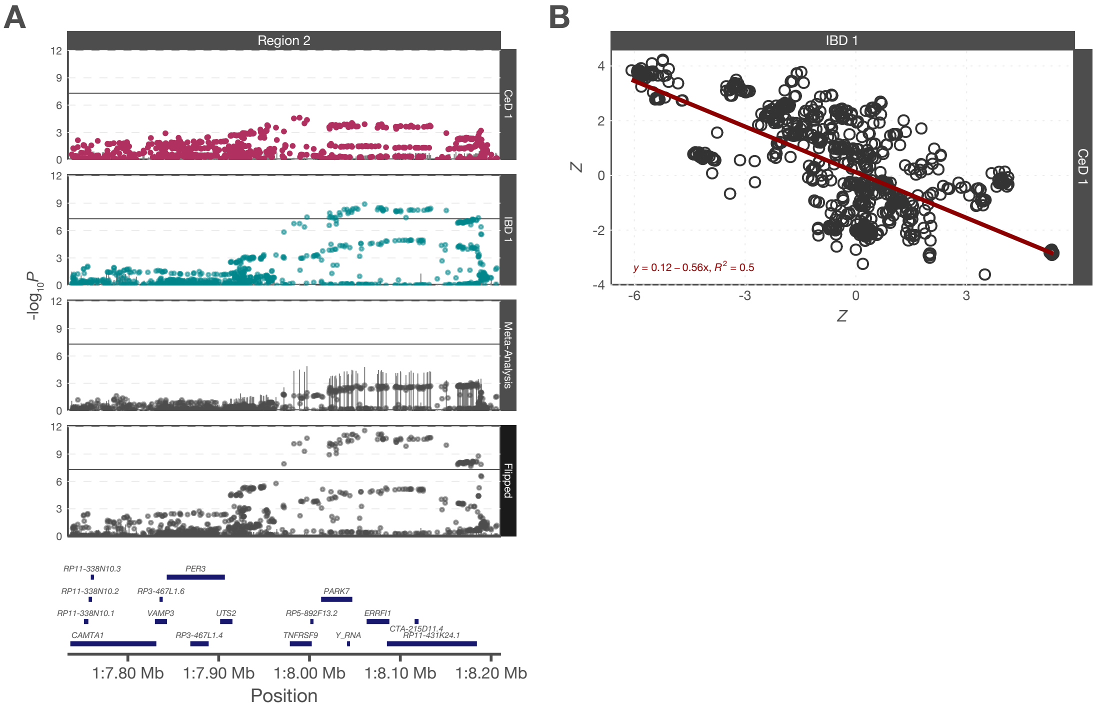


**Fig. S6:** An association signal near *TNFRSF9*, *PARK7* and *ERRFI1* exhibits opposing effects in CeD and IBD. (A) Meta-analysis of CeD and IBD association data (third panel) reduces significance (points) and increases heterogeneity (lines, Cochran's $Q$) of the association signal. (B) Regression of $Z$ scores for CeD against corresponding $Z$ scores for IBD reveals an inverse linear relationship, suggesting opposing directions of effect in the two diseases. After reversing effects for CeD and repeating meta-analysis (A, fourth panel), significance increases and heterogeneity decreases, confirming that the effects in CeD and IBD are opposed at this locus.

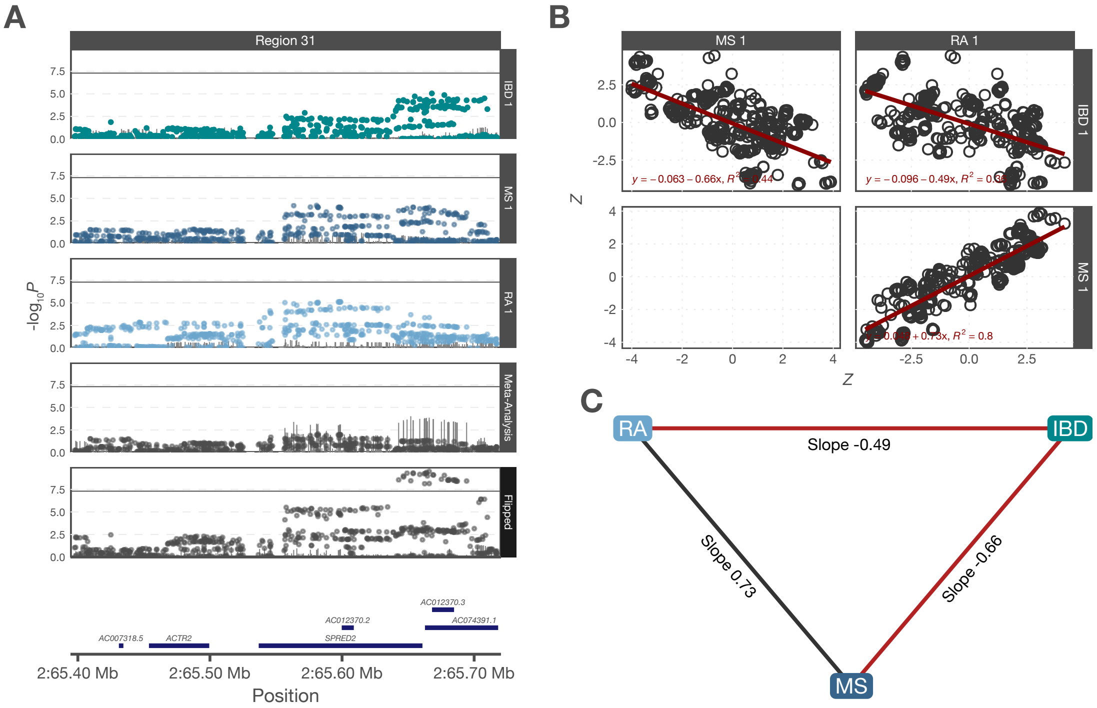


**Fig. S7:** An association signal near *SPRED2* exhibits an opposing effect in IBD compared to MS and RA. (A) Meta-analysis of IBD, MS and RA association data (fourth panel) reduces significance (points) and increases heterogeneity (lines, Cochran's *Q*) of the association signal. (B) Regression of *Z* scores for IBD against corresponding *Z* scores for MS and RA reveals inverse linear relationships, suggesting an opposing direction of effect in IBD compared to the other two diseases. By comparison, MS and RA show a positive linear relationship in their *Z* scores. After reversing effects for IBD and repeating meta-analysis (A, fifth panel), significance increases and heterogeneity decreases, confirming that the IBD effect is opposed to the effect in the other diseases (C).

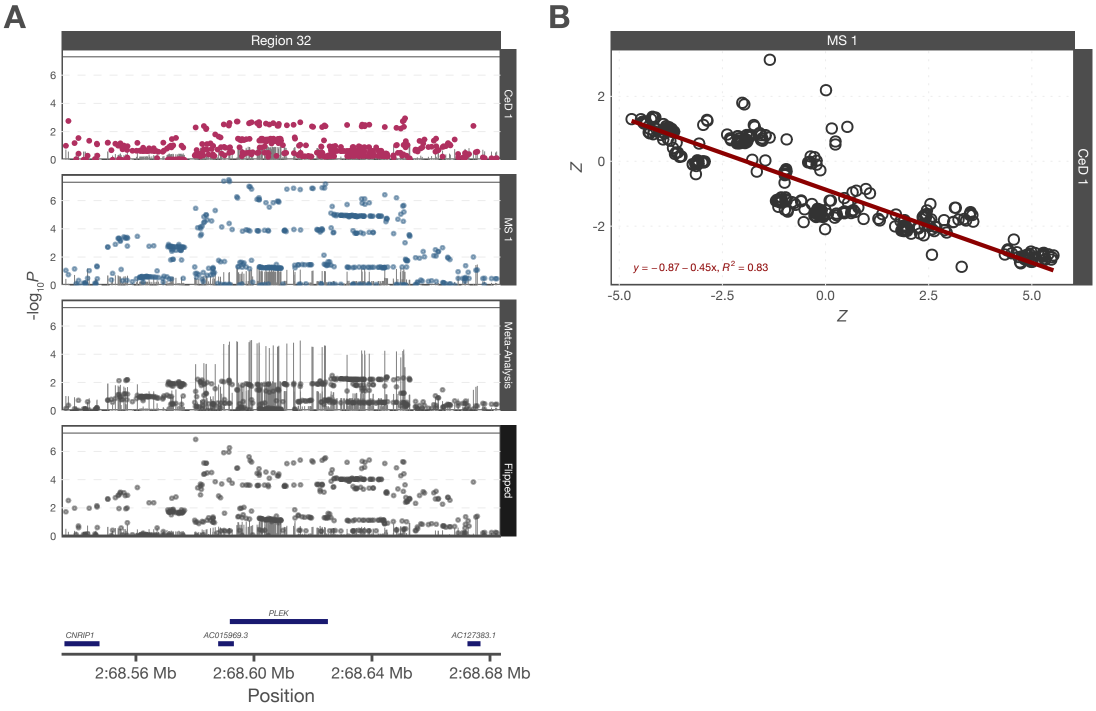


**Fig. S8:** An association signal near *PLEK* exhibits opposing effects in CeD and MS. (A) Meta-analysis of CeD and MS association data (third panel) reduces significance (points) and increases heterogeneity (lines, Cochran's *Q*) of the association signal. (B) Regression of *Z* scores for CeD against corresponding *Z* scores for MS reveals an inverse linear relationship, suggesting opposing directions of effect in the two diseases. After reversing effects for CeD and repeating meta-analysis (A, fourth panel), significance increases and heterogeneity decreases, confirming that the effects in CeD and MS are opposed at this locus.

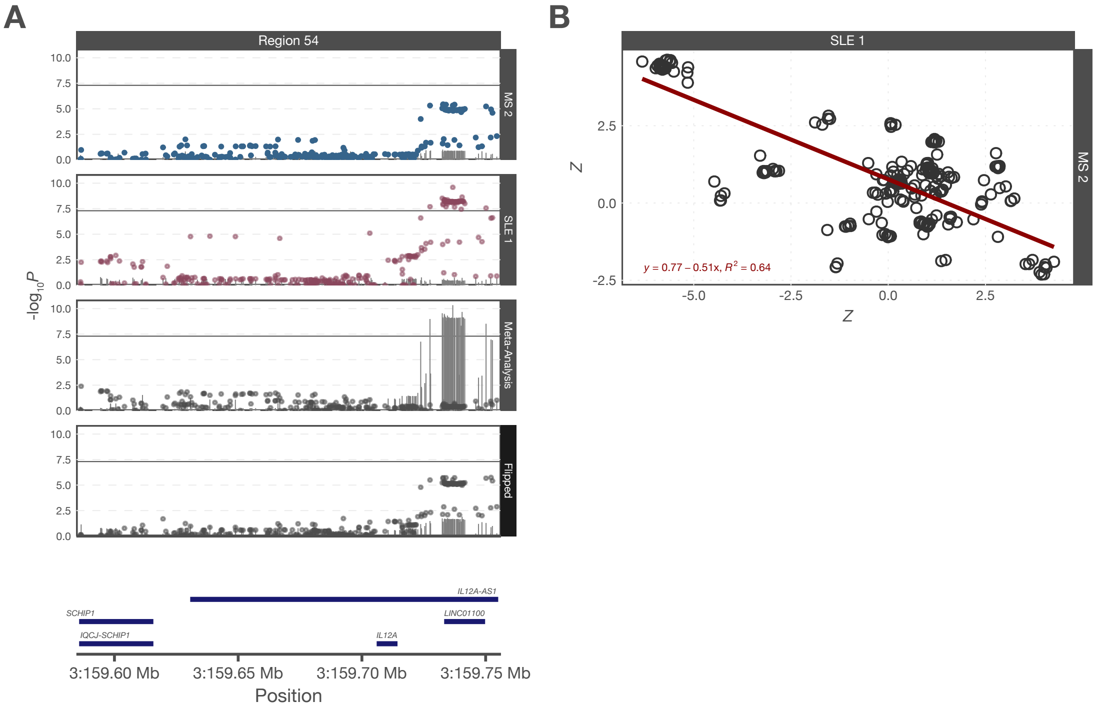


**Fig. S9:** An association signal near *IL12A* exhibits opposing effects in MS and SLE. (A) Meta-analysis of MS and SLE association data (third panel) reduces significance (points) and increases heterogeneity (lines, Cochran's *Q*) of the association signal. (B) Regression of *Z* scores for MS against corresponding *Z* scores for SLE reveals an inverse linear relationship, suggesting opposing directions of effect in the two diseases. After reversing effects for MS and repeating meta-analysis (A, fourth panel), significance increases and heterogeneity decreases, confirming that the effects in MS and SLE are opposed at this locus.

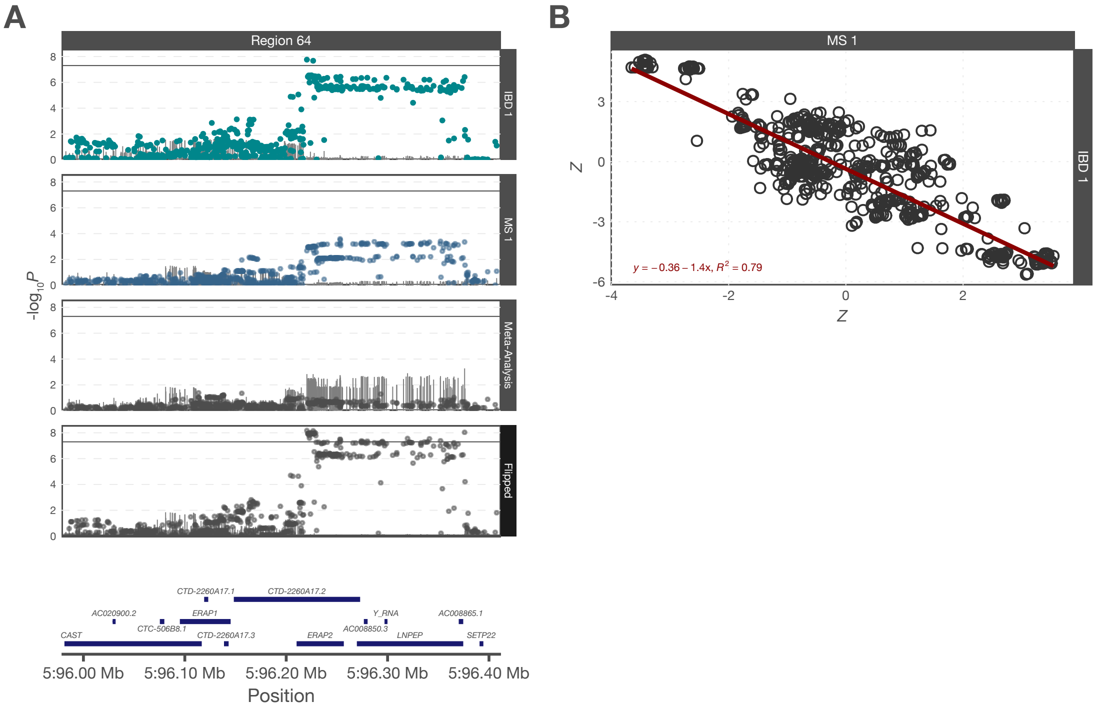


**Fig. S10:** An association signal near *ERAP2* exhibits opposing effects in IBD and MS. (A) Meta-analysis of IBD and MS association data (third panel) reduces significance (points) and increases heterogeneity (lines, Cochran's *Q*) of the association signal. (B) Regression of *Z* scores for IBD against corresponding *Z* scores for MS reveals an inverse linear relationship, suggesting opposing directions of effect in the two diseases. After reversing effects for IBD and repeating meta-analysis (A, fourth panel), significance increases and heterogeneity decreases, confirming that the effects in IBD and MS are opposed at this locus.

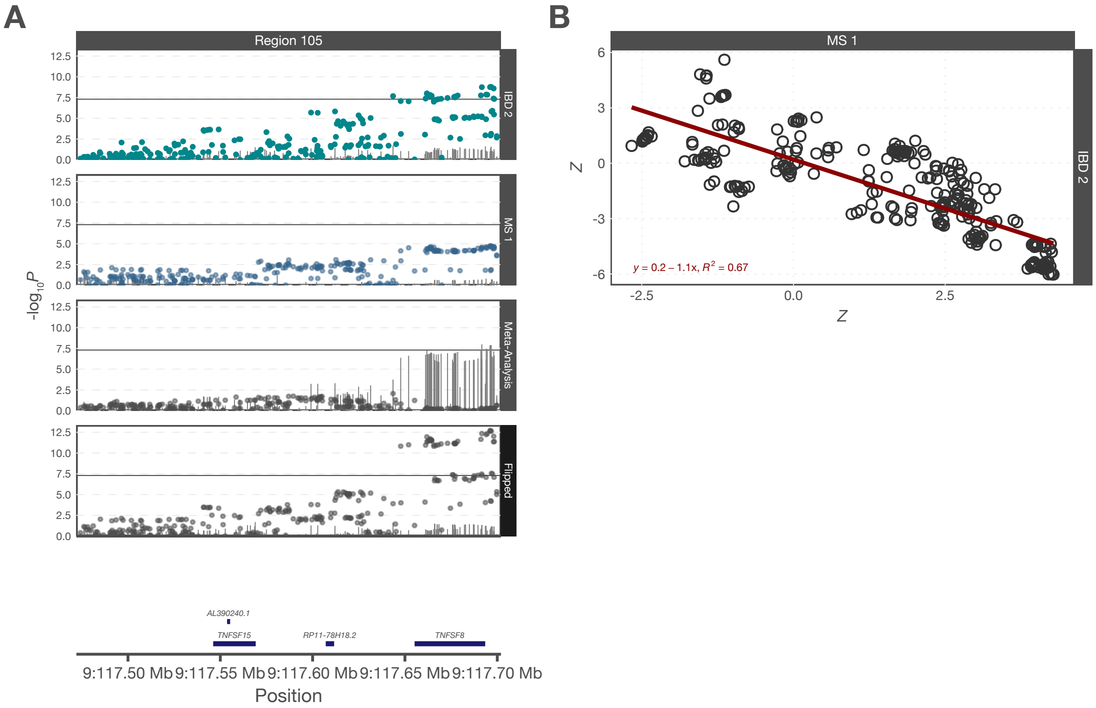


**Fig. S11:** An association signal near *TNFSF8* exhibits opposing effects in IBD and MS. (A) Meta-analysis of IBD and MS association data (third panel) reduces significance (points) and increases heterogeneity (lines, Cochran's *Q*) of the association signal. (B) Regression of *Z* scores for IBD against corresponding *Z* scores for MS reveals an inverse linear relationship, suggesting opposing directions of effect in the two diseases. After reversing effects for IBD and repeating meta-analysis (A, fourth panel), significance increases and heterogeneity decreases, confirming that the effects in IBD and MS are opposed at this locus.

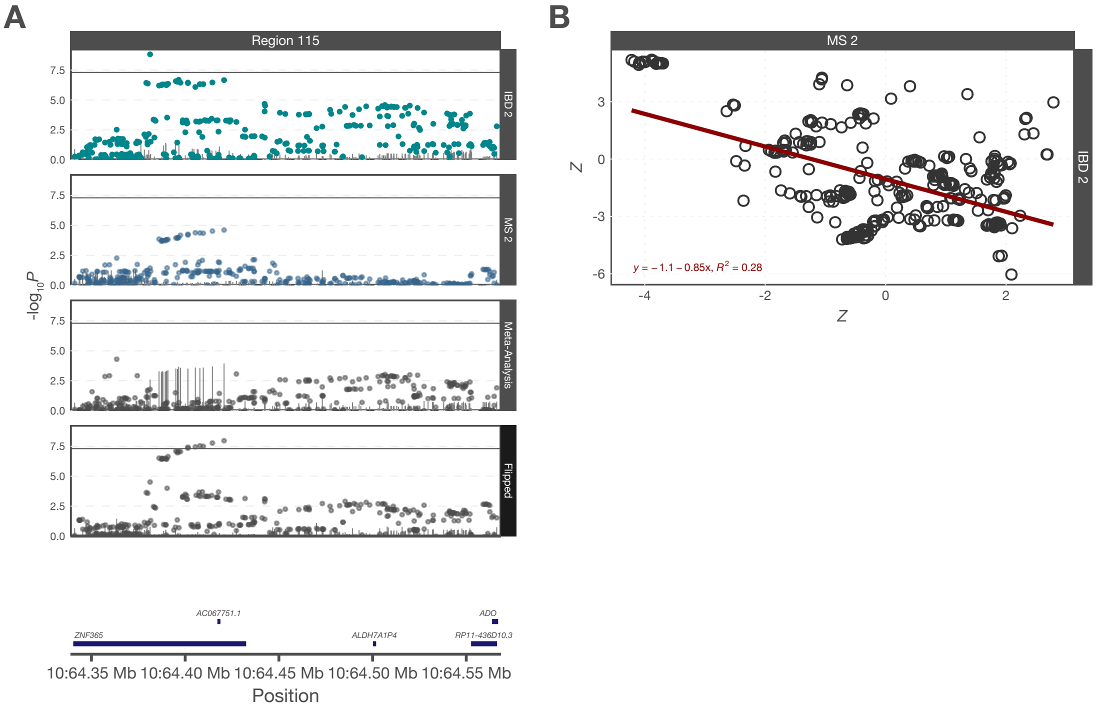


**Fig. S12:** An association signal near *ZNF365* exhibits opposing effects in IBD and MS. (A) Meta-analysis of IBD and MS association data (third panel) reduces significance (points) and increases heterogeneity (lines, Cochran's *Q*) of the association signal. (B) Regression of *Z* scores for IBD against corresponding *Z* scores for MS reveals an inverse linear relationship, suggesting opposing directions of effect in the two diseases. After reversing effects for IBD and repeating meta-analysis (A, fourth panel), significance increases and heterogeneity decreases, confirming that the effects in IBD and MS are opposed at this locus.

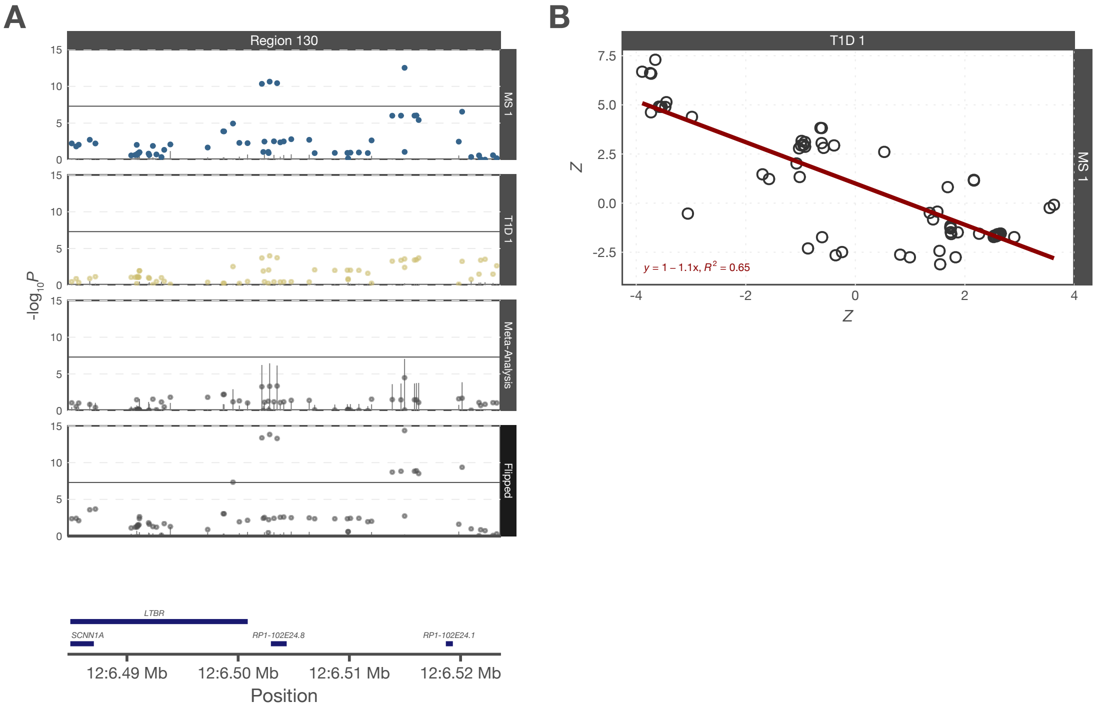


**Fig. S13:** An association signal near *LTBR* exhibits opposing effects in MS and T1D. (A) Meta-analysis of MS and T1D association data (third panel) reduces significance (points) and increases heterogeneity (lines, Cochran's *Q*) of the association signal. (B) Regression of *Z* scores for MS against corresponding *Z* scores for T1D reveals an inverse linear relationship, suggesting opposing directions of effect in the two diseases. After reversing effects for MS and repeating meta-analysis (A, fourth panel), significance increases and heterogeneity decreases, confirming that the effects in MS and T1D are opposed at this locus.

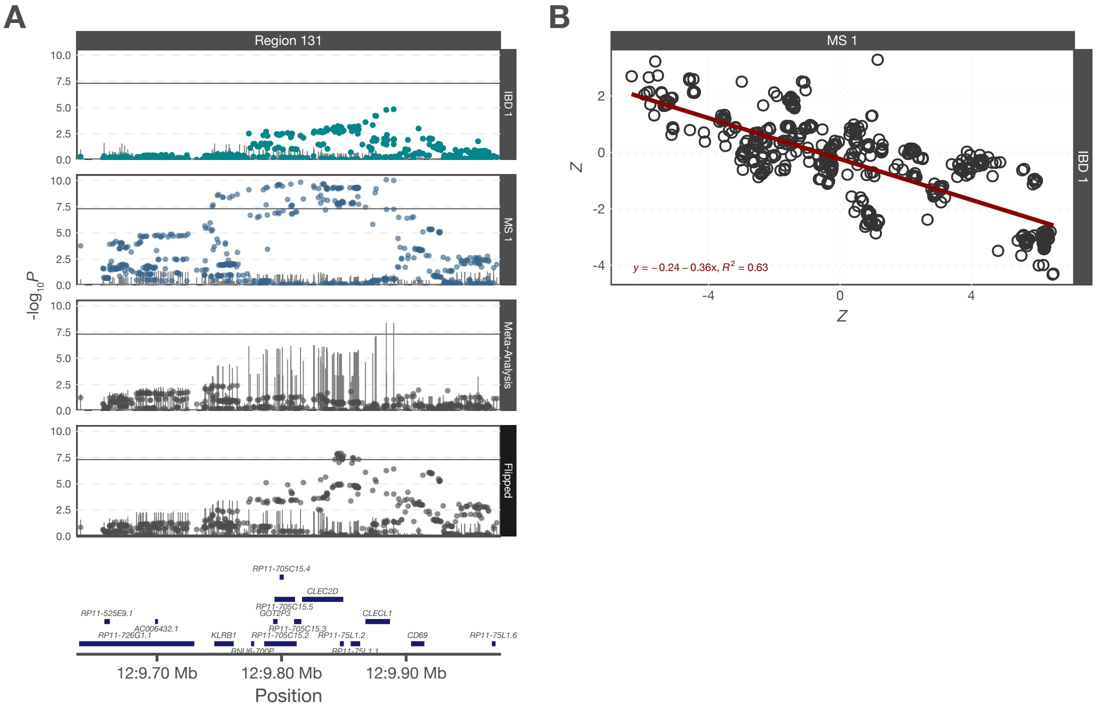


**Fig. S14:** An association signal near *CLEC2D*, *CLECL1* and *CD69* exhibits opposing effects in IBD and MS. (A) Meta-analysis of IBD and MS association data (third panel) reduces significance (points) and increases heterogeneity (lines, Cochran's *Q*) of the association signal. (B) Regression of *Z* scores for IBD against corresponding *Z* scores for MS reveals an inverse linear relationship, suggesting opposing directions of effect in the two diseases. After reversing effects for IBD and repeating meta-analysis (A, fourth panel), significance increases and heterogeneity decreases, confirming that the effects in IBD and MS are opposed at this locus.

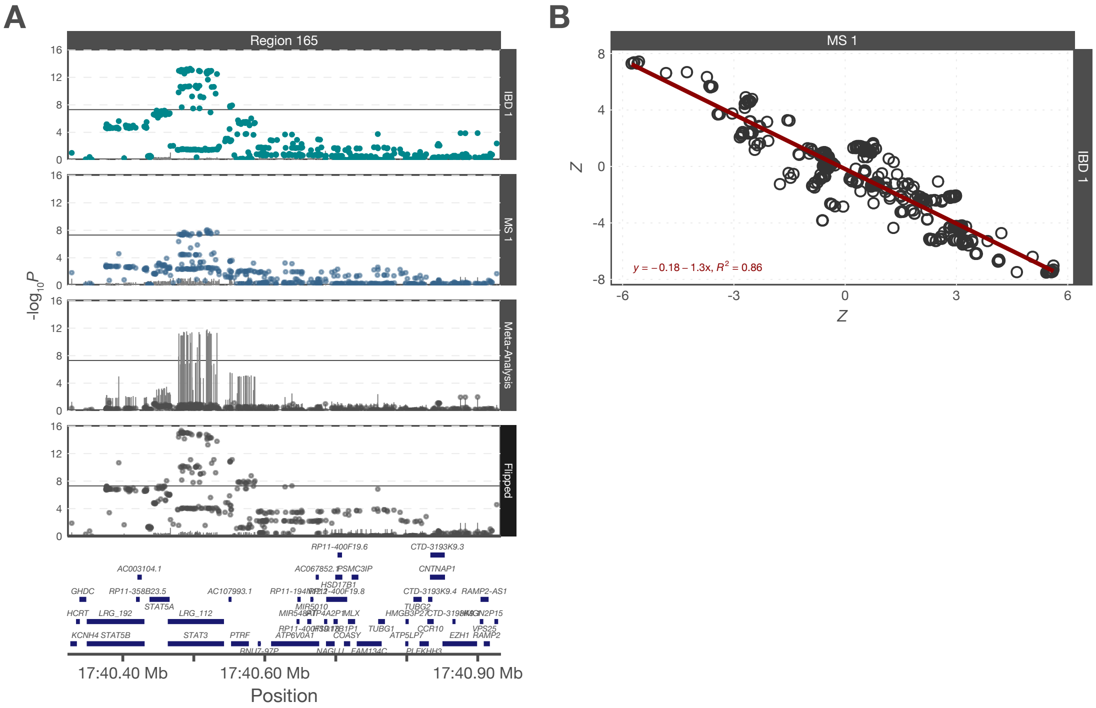


**Fig. S15:** An association signal near *STAT3* exhibits opposing effects in IBD and MS. (A) Meta-analysis of IBD and MS association data (third panel) reduces significance (points) and increases heterogeneity (lines, Cochran's *Q*) of the association signal. (B) Regression of *Z* scores for IBD against corresponding *Z* scores for MS reveals an inverse linear relationship, suggesting opposing directions of effect in the two diseases. After reversing effects for IBD and repeating meta-analysis (A, fourth panel), significance increases and heterogeneity decreases, confirming that the effects in IBD and MS are opposed at this locus.
