## Supplementary tables for "Joint analysis reveals shared autoimmune disease associations and identifies common mechanisms"

### Section 3

### Supplementary Tables

**Table S1:** GWAS and ImmunoChip studies used in the analysis. Sources for datasets used in this study. Databases and accession numbers are provided for publicly available datasets. For datasets not available in public datasets, contact details for relevant consortia are provided. NHGRI-EBI, NHGRI-EBI GWAS Catalog; dbGAP, Database of Genotypes and Phenotypes; GRASP, Genome-Wide Repository of Associations Between SNPs and Phenotypes; IBDGC, NIDDK Inflammatory Bowel Disease Genetics Consortium; IMSGC, International MS Genetics Consortium; RACI, Rheumatoid Arthritis Consortium International

| Disease | Study Type | PMID | Source | Accession | Comment |
|---|---|---|---|---|---|
| Atopic dermatitis (AtD) | GWAS | 26482879 | NHGRI-EBI | GCST003184 | |
| Allergic traits (All) | GWAS | 29083406 | NHGRI-EBI | GCST005038 | |
| Asthma (Ast) | GWAS | 32296059 | NHGRI-EBI | GCST010042 | |
| Celiac disease (CeD) | GWAS | 20190752 | NHGRI-EBI | GCST000612 | |
| Celiac disease (CeD) | ImmunoChip | 22057235 | Authors | | Summary statistics available at NHGRI-EBI; individual genotype data obtained from the authors |
| Eosinophilic granulomatosis with polyangiitis (EGPA) | GWAS | 31719529 | NHGRI-EBI | GCST009250 | |
| Selective IgA deficiency (sIgAD) | GWAS | 27723758 | NHGRI-EBI | GCST003814 | |
| Inflammatory bowel disease (IBD) | GWAS | 28067908 | NHGRI-EBI | GCST004131 | |
| Crohn's disease (CD) | GWAS | 28067908 | NHGRI-EBI | GCST004132 | |
| Ulcerative colitis (UC) | GWAS | 28067908 | NHGRI-EBI | GCST004133 | |
| Inflammatory bowel disease (IBD) | ImmunoChip | 26192919 | IBDGC | | Summary statistics available at NHGRI-EBI; individual genotype data obtained from IBDGC |
| Latent autoimmune diabetes in adults (LADA) | GWAS | 30254083 | NHGRI-EBI | GCST007245 | |
| Multiple sclerosis (MS) | GWAS | 31604244 | IMSGC | | Summary statistics available from IMSGC |
| Multiple sclerosis (MS) | ImmunoChip | 24076602 | IMSGC | | Summary statistics available from IMSGC; individual genotype data obtained from IMSGC |
| Primary biliary cirrhosis (PBC) | GWAS | 26394269 | NHGRI-EBI | GCST003129 | |
| Primary sclerosing cholangitis (PSC) | GWAS | 27992413 | NHGRI-EBI | GCST004030 | |
| Psoriasis (Ps) | GWAS | 19680446 | dbGAP | phs000019.v1.p1 | |
| Psoriatic arthritis (PsA) | GWAS | 30552173 | NHGRI-EBI | GCST007043 | |
| Rheumatoid arthritis (RA) | GWAS | 24390342 | GRASP | | NHLBI GRASP catalog, Build 2.0.0.0 |
| Rheumatoid arthritis (RA) | ImmunoChip | 23143596 | RACI | | Summary statistics available at NHGRI-EBI; individual genotype data obtained from RACI |
| Systemic lupus erythematosus (SLE) | GWAS | 26502338 | NHGRI-EBI | GCST003156 | |
| Systemic lupus erythematosus (SLE) Genentech | ImmunoChip | 28714469 | Authors | | Summary statistics available at NHGRI-EBI; individual genotype data obtained from the authors |
| Systemic lupus erythematosus (SLE) OMRF | ImmunoChip | 28135245 | Authors | | Individual genotype data obtained from the authors |
| Systemic sclerosis (SSc) | GWAS | 31672989 | NHGRI-EBI | GCST009131 | |
| Type 1 diabetes (T1D) | ImmunoChip | 25751624 | dbGAP | phs000911.v1.p1 | Individual genotype data obtained from the authors |

**Table S1:** GWAS and ImmunoChip studies used in the analysis. Sources for datasets used in this study. Databases and accession numbers are provided for publicly available datasets. For datasets not available in public datasets, contact details for relevant consortia are provided. NHGRI-EBI, NHGRI-EBI GWAS Catalog; dbGAP, Database of Genotypes and Phenotypes; GRASP, Genome-Wide Repository of Associations Between SNPs and Phenotypes; IBDGC, NIDDK Inflammatory Bowel Disease Genetics Consortium; IMSGC, International MS Genetics Consortium; RACI, Rheumatoid Arthritis Consortium International *(continued)*

| Disease | Study Type | PMID | Source | Accession | Comment |
|---|---|---|---|---|---|
| Vitiligo (Vit) | GWAS | 27723757 | NHGRI-EBI | GCST004785 | |

**Table S2:** Subjects by disease, stratum and phenotype class. Case and control counts are provided for each disease and stratum, before and after quality control filtering.

| | | Before QC | | | After QC | | |
|---|---|---|---|---|---|---|---|
| **Disease** | **Stratum** | **Cases** | **Controls** | **Unknown** | **Cases** | **Controls** | **Unknown** |
| CeD | British | 7728 | 9608 | 0 | 6903 | 8915 | 0 |
| CeD | Dutch | 946 | 150 | 119 | NA | NA | NA |
| CeD | Gosias_mystery | 1139 | 1279 | 377 | 1087 | 1144 | 288 |
| CeD | Indian | 398 | 564 | 15 | NA | NA | NA |
| CeD | Italian | 1015 | 1291 | 0 | 974 | 1225 | 0 |
| CeD | Polish | 548 | 650 | 0 | 520 | 583 | 0 |
| CeD | Romanian | 0 | 95 | 0 | NA | NA | NA |
| CeD | Spanish | 522 | 437 | 0 | 482 | 349 | 0 |
| CeD | Unknown | 0 | 0 | 82 | NA | NA | NA |
| IBD | Australia | 1737 | 533 | 0 | 1681 | 507 | 0 |
| IBD | Belgium | 4347 | 1734 | 20 | 4132 | 1621 | 19 |
| IBD | Denmark | 238 | 90 | 0 | 235 | 90 | 0 |
| IBD | Germany | 3611 | 6598 | 0 | 3392 | 6467 | 0 |
| IBD | IMSGC | 0 | 5789 | 0 | NA | NA | NA |
| IBD | Iran | 302 | 111 | 0 | NA | NA | NA |
| IBD | Italy | 2203 | 1797 | 0 | 2083 | 1710 | 0 |
| IBD | Lithuania-Baltic | 450 | 283 | 0 | 433 | 281 | 0 |
| IBD | Netherlands | 2272 | 1753 | 10 | 2140 | 1716 | 8 |
| IBD | New_Zealand | 1234 | 498 | 0 | 1096 | 481 | 0 |
| IBD | Norway | 435 | 372 | 0 | 420 | 369 | 0 |
| IBD | Slovenia | 250 | 233 | 1 | 230 | 222 | 1 |
| IBD | Spain | 279 | 291 | 0 | 269 | 285 | 0 |
| IBD | Sweden | 1065 | 2409 | 0 | 1014 | 2358 | 0 |
| IBD | UK | 2907 | 0 | 173 | NA | NA | NA |
| IBD | Unknown | 5401 | 10477 | 921 | 5220 | 10016 | 886 |
| IBD | USA-Canada | 8280 | 1816 | 53 | 7632 | 1671 | 51 |
| MS | AUSNZ | 246 | 935 | 0 | 243 | 923 | 0 |
| MS | Belgium | 301 | 1675 | 0 | 297 | 1652 | 0 |
| MS | Denmark | 740 | 831 | 0 | 728 | 822 | 0 |
| MS | Finland | 221 | 486 | 0 | 219 | 481 | 0 |
| MS | France | 385 | 353 | 0 | 378 | 346 | 0 |
| MS | Germany | 2582 | 5533 | 0 | 2548 | 5468 | 0 |
| MS | Italy | 957 | 1255 | 0 | 948 | 1237 | 0 |
| MS | Norway | 894 | 674 | 0 | 879 | 665 | 0 |
| MS | Sweden | 2153 | 2331 | 0 | 2123 | 2298 | 0 |
| MS | UK | 4313 | 4408 | 0 | 4247 | 4359 | 0 |
| MS | Unknown | 16 | 120 | 0 | NA | NA | NA |
| MS | US | 1690 | 5490 | 0 | 1661 | 5402 | 0 |
| RA | ES | 907 | 447 | 0 | 802 | 417 | 0 |
| RA | NL | 702 | 2079 | 0 | 650 | 1993 | 0 |
| RA | SE-E | 2962 | 2098 | 0 | 2775 | 1990 | 0 |
| RA | SE-U | 1014 | 1018 | 0 | 887 | 958 | 0 |
| RA | UK | 4189 | 8900 | 0 | 3903 | 8418 | 0 |
| RA | US | 3468 | 3820 | 0 | 3111 | 2200 | 0 |
| SLE | sle_g.AA | 455 | 824 | 0 | NA | NA | NA |
| SLE | sle_g.EA | 3834 | 10663 | 3 | 3299 | 9602 | 3 |
| SLE | sle_g.Others | 127 | 127 | 0 | NA | NA | NA |
| SLE | sle_o | 2563 | 2054 | 0 | 1149 | 1177 | 0 |
| T1D | GRID | 6670 | 9416 | 0 | 6551 | 9152 | 0 |
| T1D | ASP | 5571 | 5220 | 0 | 5289 | 5003 | 0 |

**Table S3:** eQTLs identified at disease level and after cross-disease meta-analysis. Shared disease susceptibility—eQTL associations are indicated. Conditioning SNPs for the eQTL data are indicated where appropriate. Where an eQTL is identified at the disease level and also in cross-disease meta-analysis, this is indicated as "Stable eQTL." eQTLs that are newly identified after cross-disease meta-analysis are labelled "New eQTL."

| Disease | Loci Shared | Loci Not Shared | Loci Not Analyzed |
|---|---|---|---|
| CeD | 17 | 6 | 13 |
| IBD | 25 | 25 | 60 |
| MS | 23 | 18 | 23 |
| RA | 20 | 7 | 20 |
| SLE | 13 | 9 | 27 |
| T1D | 12 | 11 | 24 |
